# MyGeneRisk Colon: A Web-Based Tool for Personalized Colorectal Cancer Risk Prediction Based on Genetics and Lifestyle

**DOI:** 10.64898/2026.04.03.26349669

**Authors:** Jiayin Zheng, Robert S Steinfelder, Hang Yin, Conghui Qu, Minta Thomas, Sushma S Thomas, Cynthia Andrews, Bianca Augusto, Douglas C Corley, Jeffrey K Lee, Sonja I Berndt, Andrew T Chan, Stephen J Chanock, Chris Gignoux, Shauna R Goldberg, Christopher A Haiman, Jeroen R Huyghe, Motoki Iwasaki, Loic Le Marchand, Soo Chin Lee, Johana Melendez, Ivan Mesa, Shuji Ogino, Viviam Sifontes, Caroline Y Um, Kala Visvanathan, Larissa L White, Andrea Williams, Waverly Willis, Alicja Wolk, Taiki Yamaji, Susan T Vadaparampil, Gail P Jarvik, Andrea N Burnett-Hartman, Roger L Milne, Elizabeth A Platz, Jane C Figueiredo, Wei Zheng, Robert J MacInnis, Julie R Palmer, Stephanie L Schmit, Iris Landorp-Vogelaar, Ulrike Peters, Li Hsu

## Abstract

Colorectal cancer (CRC) is a leading cause of cancer-related death, with incidence rising substantially among individuals under 50 years of age. Polygenic risk scores (PRS) hold promise for identifying high-risk individuals; when combined with lifestyle factors, they substantially improve prediction accuracy compared with models based on lifestyle factors alone. However, few clinical tools currently exist that facilitate this integrated, PRS-enhanced risk assessment. To bridge this gap, we developed ***MyGeneRisk Colo****n,* a publicly accessible web portal that delivers individualized CRC risk prediction by incorporating genetic, demographic, family history, and lifestyle factors. This paper details the development of the underlying risk prediction model, the portal’s architecture and data security, our reporting framework, and engagement with a community advisory panel. Designed as a user-friendly platform, *MyGeneRisk Colon* aims to effectively communicate personalized CRC risk profiles and educate users and healthcare providers about prevention strategies.

## Introduction

Colorectal cancer (CRC) remains a leading cause of cancer-related mortality, with a particularly alarming increase in incidence among people under the age of 50.^1^ Screening, such as colonoscopy, substantially reduces the risk of developing CRC and dying from it. Although screening recommendations in the US have been lowered in recent years to age 45^2^, more than half of early-onset cases are diagnosed before the age of 45,^3^ highlighting the need for improved risk stratification to inform personalized prevention strategies.

Effective prevention of CRC relies on accurate risk assessment; however, traditional risk factors, such as family history, are absent in over 80% of CRC cases. The discovery of over 200 common genetic risk loci for CRC^4–17^ has spurred the development of polygenic risk scores (PRS), which aggregate the small effects of individual genetic susceptibility variants^4–10,18^. PRS models that include genome-wide variants beyond the established loci have demonstrated improved accuracy in predicting CRC risk.^16^ Furthermore, predictive performance is significantly enhanced when PRS are integrated with lifestyle factors, such as obesity, smoking, red meat intake, and alcohol consumption.^19^ The multifactorial nature of CRC presents an opportunity to develop comprehensive risk prediction models that synergize genetic and lifestyle determinants.

Despite this scientific progress,^16,20–26^, readily accessible clinical tools for PRS-enhanced CRC risk assessment remain limited. Most existing models were developed predominantly in populations of European ancestry, rendering them less accurate and transferable across diverse racial and ethnic groups, thereby limiting their broader clinical utility. In addition, a persistent gap exists between scientific advances in risk modeling and their translation into user-friendly, publicly accessible tools. This gap is often worsened by limited community engagement.

Without meaningful community input, scientists may overlook public concerns, ethical implications, and usability of risk models, hindering the translation of promising scientific advances into real-world applications.^27^

To bridge these critical gaps, we developed MyGeneRisk Colon. This user-friendly web portal combines self-reported lifestyle information with PRS calculated from users’ raw genotyping data, which individuals can download directly from direct-to-consumer genetic testing (DTC-GT) services (e.g., Ancestry.com and 23andMe). We view MyGeneRisk Colon as a proof of principle to demonstrate that complex genome-wide PRS calculations and imputation can be performed in real-time to generate clear, actionable risk reports. A key technological feature is the platform’s ability to impute millions of genetic variants based on the uploaded DTC-GT genetic data and calculate a genome-wide PRS in real-time, completing the analysis in minutes while the user answers lifestyle questions.

The underlying prediction model was rigorously developed based on large-scale, diverse studies. In particular, we derived relative risk estimates for lifestyle factors from harmonized individual participant data across sixteen diverse prospective cohorts over 670,000 individuals (∼17,000 CRC cases) and calculated absolute risk using data from national cancer statistics. A genome-wide PRS derived from 100,204 CRC cases and 154,587 controls was incorporated into the model. The PRS was independently validated in Asian, Black or African American, Hispanic, and non-Hispanic White populations.^16^ Although the platform is currently designed for the growing population of individuals who already have their DTC-GT data, it establishes a technical and reporting infrastructure that can be integrated into electronic health records for routine clinical care and expanded to other complex diseases. This paper details the model development, portal architecture, data security measures, and community engagement process. Ultimately, this user-friendly platform aims to effectively communicate personalized CRC risk, empowering individuals and healthcare providers to make informed, evidence-based decisions about prevention.

## Results

### Study Sample

As our goal was to develop risk prediction models for estimating the risk of developing CRC, we included cohort studies or cohort-based nested case-control studies with clinical and epidemiological data. In total, we included 16 cohorts representing diverse populations with 673,265 individuals (17,355 CRC cases): Black Women’s Health Study (BWHS), Campaign Against Cancer and Heart Disease (CLUE II), Cancer Prevention Study II (CPS II), Health Professionals Follow-up Study (HPFS), The Japan Public Health Center-based prospective Study (JPHC), Melbourne Collaborative Cohort Study (MCCS), Multiethnic Cohort Study (MEC), Nurses’ Health Study (NHS), Nurses’ Health Study II (NHSII), Physician’s Health Study (PHS), Prostate, Lung, Colorectal and Ovarian Cancer Screening Trail (PLCO), Southern Community Cohort Study (SCCS), Swedish Mammography Cohort and Swedish Men Cohort (SMC_COSM), UK Biobank (UKB), VITamins And Lifestyle (VITAL), and Women’s Health Initiative (WHI). Descriptive statistics of these studies are shown in **Table 1**, and further details are provided in the supplemental materials.

**Table 1.** Description of studies in model development.

| Study | Sample Size | CRC | Age | Female | Asian* | Black/<br>African American* | Hispanic | White* | Other* |
| --- | --- | --- | --- | --- | --- | --- | --- | --- | --- |
|  |  |  |  |  | 19597<br>(1096)<sup>&</sup> | 12877<br>(880)<sup>&</sup> | 3083<br>(66)<sup>&</sup> | 622337<br>(15078)<sup>&</sup> | 15371<br>(235)<sup>&</sup> |
| *Nested Case-control studies* |  |  |  |  |  |  |  |  |  |
| BWHS | 615 | 30.9% | 25-69 | 100% | 0.0% | 98.9% | 1.1% | 0.0% | 0.0% |
| CLUE II | 607 | 49.1% | 31-89 | 54% | 0.0% | 2.0% | 0.0% | 97.5% | 0.5% |
| CPSII | 4799 | 48.9% | 48-85 | 48% | 0.4% | 0.9% | 0.0% | 97.9% | 0.8% |
| HPFS | 1908 | 48.8% | 41-94 | 0% | 0.8% | 0.9% | 0.0% | 92.7% | 5.5% |
| JPHC | 1265 | 48.1% | 40-69 | 49% | 100.0% | 0.0% | 0.0% | 0.0% | 0.0% |
| MCCS | 2314 | 50.6% | 38-76 | 49% | 0.0% | 0.0% | 0.0% | 100.0% | 0.0% |
| MEC | 1563 | 48.7% | 45-77 | 48% | 28.9% | 14.3% | 0.0% | 56.7% | 0.0% |
| NHS | 2945 | 48.0% | 35-89 | 100% | 0.3% | 1.2% | 1.1% | 97.4% | 0.1% |
| NHSII | 289 | 54.3% | 29-64 | 100% | 0.7% | 0.7% | 1.4% | 97.2% | 0.0% |
| PHS | 900 | 44.4% | 40-85 | 0% | 2.1% | 0.4% | 1.2% | 84.4% | 11.8% |
| SCCS | 730 | 46.7% | 40-79 | 62% | 0.0% | 100.0% | 0.0% | 0.0% | 0.0% |
| SMC_COSM | 1626 | 37.6% | 45-83 | 39% | 0.0% | 0.0% | 0.0% | 100.0% | 0.0% |
| VITAL | 741 | 49.5% | 50-76 | 45% | 1.9% | 0.9% | 0.4% | 92.3% | 4.5% |
| WHI | 4160 | 49.2% | 50-79 | 100% | 2.4% | 0.0% | 0.0% | 97.6% | 0.0% |
| *Cohort studies* |  |  |  |  |  |  |  |  |  |
| PLCO | 149395 | 2.0% | 42-78 | 51% | 4.2% | 5.10% | 2.00% | 88.40% | 0.20% |
| UKB | 499408 | 0.6% | 37-73 | 54% | 2.3% | 0.70% | 0.00% | 94.10% | 3.00% |

\*Non-Hispanic. <sup>&</sup> Sample size (# of cases) for each racial and ethnic group.

### Lifestyle Factors

We selected putative or known risk factors associated with CRC risk, including anthropometric traits, behavioral factors, diet, pharmacological factors, and medical history. Specifically, we included height, body mass index, smoking status, smoking pack-years, alcohol consumption, dietary factors (intake of calcium supplements, red meat and, fruit), regular aspirin use, regular non-aspirin nonsteroidal anti-inflammatory drug (NSAIDS) use, regular use of postmenopausal hormones, history of type 2 diabetes mellitus, history of endoscopy (colonoscopy or sigmoidoscopy), and first-degree family history of CRC. To distinguish these from genetic factors, we refer to them as ’lifestyle factors’ throughout the paper for simplicity. Due to the high correlation between intake of red meat and processed meat, and between intake of vegetables and fruit, we included only red meat and fruit intake in the risk model. Across studies, risk factor information was collected by in-person interviews and/or self-administered structured questionnaires, as detailed previously^19^ and in the Supplemental Materials. All factors were collected at the study entry or blood collection. To harmonize each risk factor across studies, we employed a multistep data harmonization procedure, described in detail by Hutter et al^28^. A brief overview of the harmonization process and individual risk factors is provided in the Harmonization of Lifestyle Factors section of the Supplemental Materials.

**Figures 1 and 2** show stackplots or boxplots of risk factors by study and sex. **Table S1a** in the Supplemental Materials summarizes the descriptive characteristics of lifestyle risk factors. For continuous variables, we applied sex-specific winsorization to ensure robustness against outliers. Height and BMI were winsorized at the 2.5th and 97.5th percentiles, while other continuous risk factors were winsorized at the 95th percentile. These percentiles were calculated using pooled data from all studies, comprising 180 imputed datasets, as described in the next section. **Table S1b and S1c** provide information on missingness and the winsorization cutoffs, respectively.

**Figure 1:**
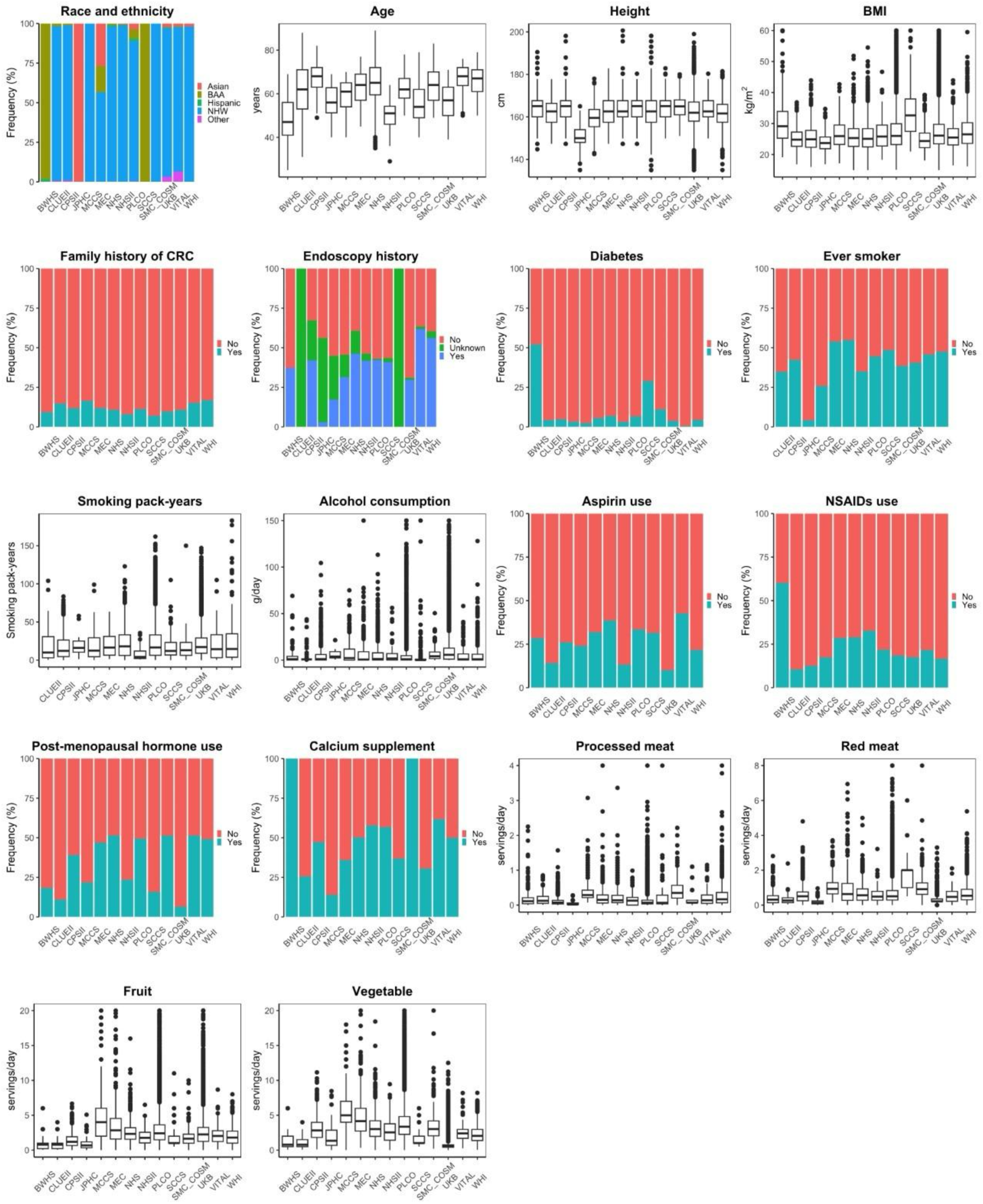
Distribution of 18 lifestyle factors in 14 studies among females. Stacked plots are used for categorical variables, and bar plots are used for continuous variables. For cohort-based nested case-control studies, distributions are presented for controls only. Abbreviations: BAA, Black or African American; NHW, non-Hispanic White; BMI, body mass index; NSAIDs, nonsteroidal anti-inflammatory drugs. Some variables were systematically missing in certain studies and are therefore not displayed. An exception is endoscopy history, for which an ‘unknown’ category was included to represent participants with missing endoscopic status.

**Figure 2:**
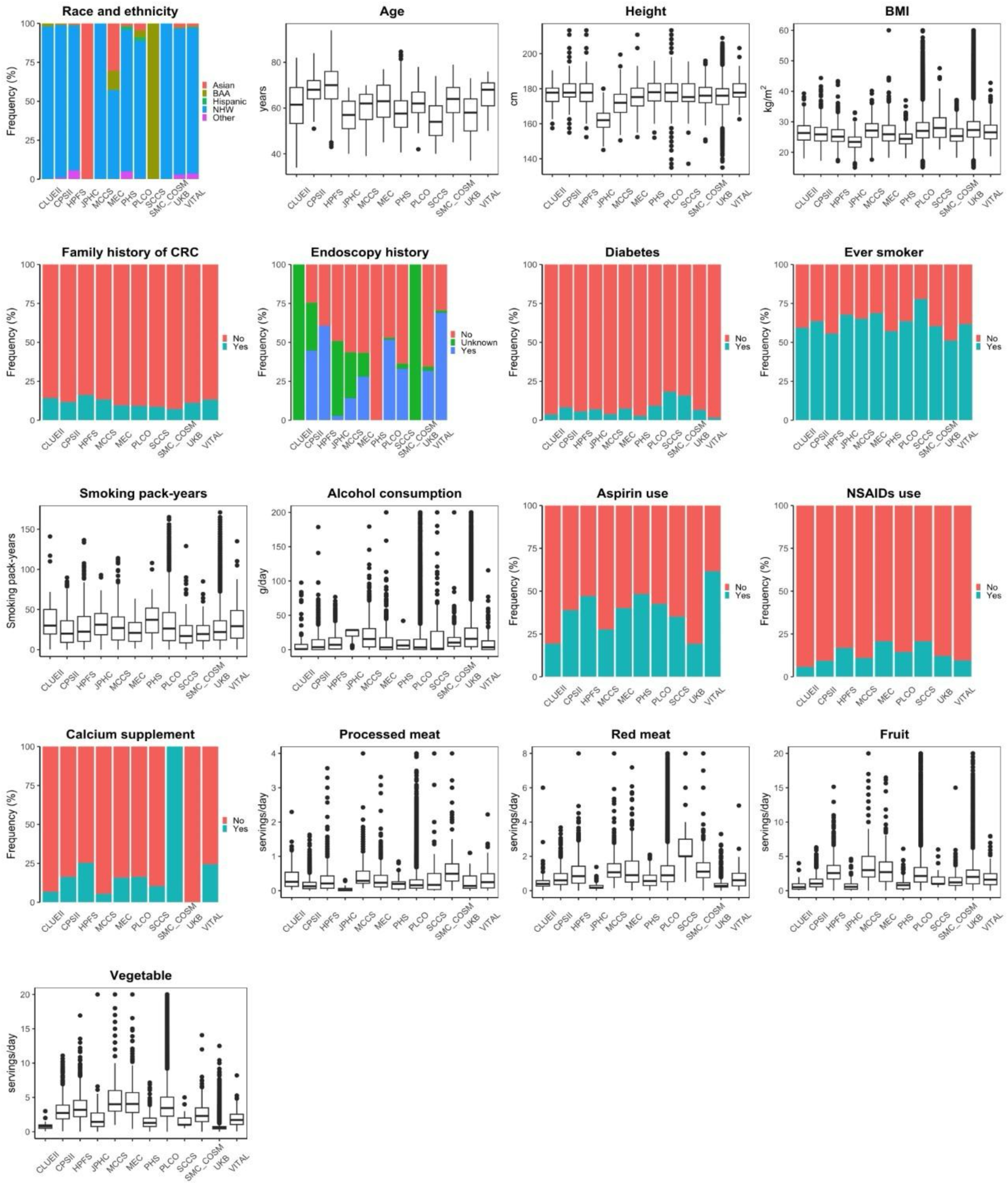
Distribution of 17 lifestyle factors in 12 studies among males. Stacked plots are used for categorical variables, and bar plots are used for continuous variables. For cohort-based nested case-control studies, distributions are presented for controls only. Abbreviations: BAA, Black or African American; NHW, non-Hispanic White; BMI, body mass index; NSAIDs, nonsteroidal anti-inflammatory drugs. Some variables were systematically missing in certain studies and are therefore not displayed. An exception is endoscopy history, for which an ‘unknown’ category was included to represent participants with missing endoscopic status.

### Risk Prediction Model Development

To estimate the association of lifestyle factors with CRC risk, we conducted an individual participant data meta-analysis using a two-step approach. In the first step, for each study, we performed multiple imputation to account for missing data, fit regression models to each imputed dataset, and then used Rubin’s rules to pool the HR estimates. In the second step, we performed multivariate random-effects meta-analysis to synthesize the study-specific HRs, allowing for between-study heterogeneity by incorporating random study effects.

**Table 2** summarizes the association of lifestyle factors with CRC, and **Figures S1 and S2** show the forest plots of HR for each risk factor. Generally, being tall, being obese, having a positive first-degree family history of CRC, having diabetes, smoking cigarettes, drinking alcohol, and having higher red meat intake were associated with increased CRC risk, while having had endoscopy, using aspirin or NSAIDS, and having a higher intake of calcium and fruit were associated with reduced risk. For females, post-menopausal hormone use was also associated with lower CRC risk.

**Table 2.** Summary of the association of lifestyle factors with CRC by sex.

|  | Female |  | Male |  |
| --- | --- | --- | --- | --- |
|  | Hazard Ratio<br>(95% CI) | Prediction<br>Interval | Hazard Ratio<br>(95% CI) | Prediction<br>Interval |
| Height, per 10 cm | 1.12 (1.07, 1.17) | (1.07, 1.17) | 1.05 (1.01, 1.10) | (1.01, 1.10) |
| BMI, per 5 kg/m<sup>2</sup> | 1.08 (1.03, 1.12) | (0.99, 1.17) | 1.16 (1.10, 1.22) | (1.05, 1.27) |
| Family history of CRC, yes | 1.28 (1.18, 1.38) | (1.18, 1.38) | 1.28 (1.18, 1.40) | (1.18, 1.40) |
| Endoscopy history, missing versus yes | 1.25 (0.95, 1.64) | (0.61, 2.57) | 1.50 (1.02, 2.19) | (0.57, 3.95) |
| Endoscopy history, no versus yes | 1.45 (1.25, 1.68) | (0.96, 2.20) | 1.56 (1.34, 1.80) | (1.10, 2.21) |
| Diabetes, yes | 1.31 (1.17, 1.46) | (1.17, 1.46) | 1.24 (1.12, 1.37) | (1.12, 1.37) |
| Smoking status, ever smoker | 1.02 (0.95, 1.11) | (0.95, 1.11) | 1.04 (0.96, 1.12) | (0.96, 1.12) |
| Smoking, per 10 pack-years | 1.09 (1.05, 1.13) | (1.02, 1.17) | 1.06 (1.04, 1.08) | (1.04, 1.08) |
| Alcohol consumption, per 14 g/day | 1.01 (0.94, 1.09) | (0.88, 1.17) | 1.08 (1.04, 1.12) | (0.99, 1.17) |
| Aspirin use, yes | 0.91 (0.81, 1.01) | (0.69, 1.20) | 0.88 (0.83, 0.94) | (0.83, 0.94) |
| NSAIDs (non-aspirin) use, yes | 0.84 (0.76, 0.93) | (0.68, 1.04) | 0.80 (0.73, 0.88) | (0.73, 0.88) |
| Post-menopausal hormone use, yes | 0.78 (0.73, 0.83) | (0.73, 0.83) | - | - |
| Calcium supplement, yes | 0.92 (0.86, 0.98) | (0.82, 1.03) | 0.88 (0.79, 0.98) | (0.79, 0.98) |
| Red meat, servings/day | 1.01 (0.92, 1.12) | (0.92, 1.12) | 1.11 (0.99, 1.24) | (0.87, 1.40) |
| Fruit, servings/day | 0.99 (0.97, 1.01) | (0.97, 1.01) | 1.00 (0.97, 1.02) | (0.96, 1.04) |

CI: confidence interval

We report both 95% confidence intervals and prediction intervals for completeness; however, for prospective risk assessment in the web tool, we used the 95% confidence intervals. We observed that the prediction intervals were substantially wider for endoscopy-related variables, reflecting high heterogeneity likely driven by historical changes in screening guidelines over the decades. As this temporal variation does not represent present-day screening standards widely adopted by the clinical community, prediction intervals may exaggerate the level of heterogeneity; therefore, we opted to use confidence intervals to quantify uncertainty in risk estimation.

To model genetic susceptibility, we incorporated the Asian-Euro polygenic risk score (PRS) developed based on 69,175 individuals of Asian ancestry (21,731 cases; 47,444 controls) and 185,616 individuals of European ancestry (78,473 cases; 107,143 controls) and validated in independent datasets across four major self-reported groups: Asian, Black or African American, Hispanic, and non-Hispanic White ^16^. As the study samples used to derive the relative risk estimates of lifestyle factors were also included in the PRS training dataset, directly estimating PRS effect sizes within our data would introduce overfitting and inflate risk estimates. To avoid this, we used the odds ratio estimates (**Table 3**) derived from independent validation datasets reported in Thomas et al. (2023) ^16^, stratified by self-reported racial and ethnic groups. Odds ratio is also estimated for all by combining all racial and ethnic groups using the inverse-variance weighted estimator. This estimate is used if an individual does not provide their race and ethnicity information. In that study, the PRS was strongly associated with an increased risk of CRC across all groups, with similar odds ratio estimates for the Asian, Hispanic, and non-Hispanic White groups, and attenuated estimates for the Black or African American group. This attenuation is consistent with known challenges in cross-ancestry prediction, driven by both the historical underrepresentation of African ancestry in GWAS and the distinct genetic architecture of African populations, characterized by shorter linkage disequilibrium blocks and greater genetic diversity.

**Table 3.** Odds ratios for PRS per standard deviation increase, stratified by four racial and ethnic groups, based on a previous publication.^16^.

| Racial and ethnic group | Cases/Controls | Odds Ratio (95% CI)* |
| --- | --- | --- |
| Asian | 2,420/9,605 | 1.64 (1.55, 1.74) |
| Black or African American | 1,954/11,869 | 1.39 (1.31, 1.47) |
| Hispanic | 1,681/8,696 | 1.62 (1.51, 1.73) |
| Non-Hispanic White | 3,651/115,105 | 1.67 (1.60, 1.75) |
| All | 9,706/145,275 | 1.59 (1.55, 1.63) |

\*CI: confidence interval; PRS were recalibrated and standardized using data from the 1000 Genomes Project to enable consistent interpretation of odds ratios across populations.

To estimate the absolute risk, we first estimated attributable risks (ARs) for lifestyle factors and PRS using novel sampling-based and analytical methods. We then obtained the sex- and race- and ethnicity-specific baseline hazard rates by multiplying the Surveillance, Epidemiology, and End Results (SEER) CRC incidence rates by one minus the ARs.^29^ Next, we combined the HR estimates for PRS and lifestyle factors with baseline age-specific cancer hazard rates to calculate absolute risk, accounting for competing risks of mortality from causes other than CRC. Specifically, we calculated the probability of developing CRC over a prespecified time interval (e.g., 5, 10, or 20 years), given an individual’s current age and risk factor profile.

We provided three options for generating CRC absolute risk estimates: PRS only, lifestyle factors only, or a combination of PRS and lifestyle factors. An example is shown in the bottom panel of **Figure 3**. The genotyping file was obtained from the Personal Genome Project (Church 2005, ID: hu33515), and the lifestyle factor inputs were described at the end of the risk reports (Supplemental Materials). In this example, the individual’s PRS indicates a higher-than-average CRC risk, whereas their lifestyle profile suggests a lower risk; and when both PRS and lifestyle factors are considered jointly, the estimated risk is only slightly above average. Notably, the 95% confidence intervals are very narrow for the PRS-based absolute risk estimate but moderately wide for the lifestyle factor-based estimates, reflecting somewhat greater uncertainty associated with the lifestyle factors.

**Figure 3:**
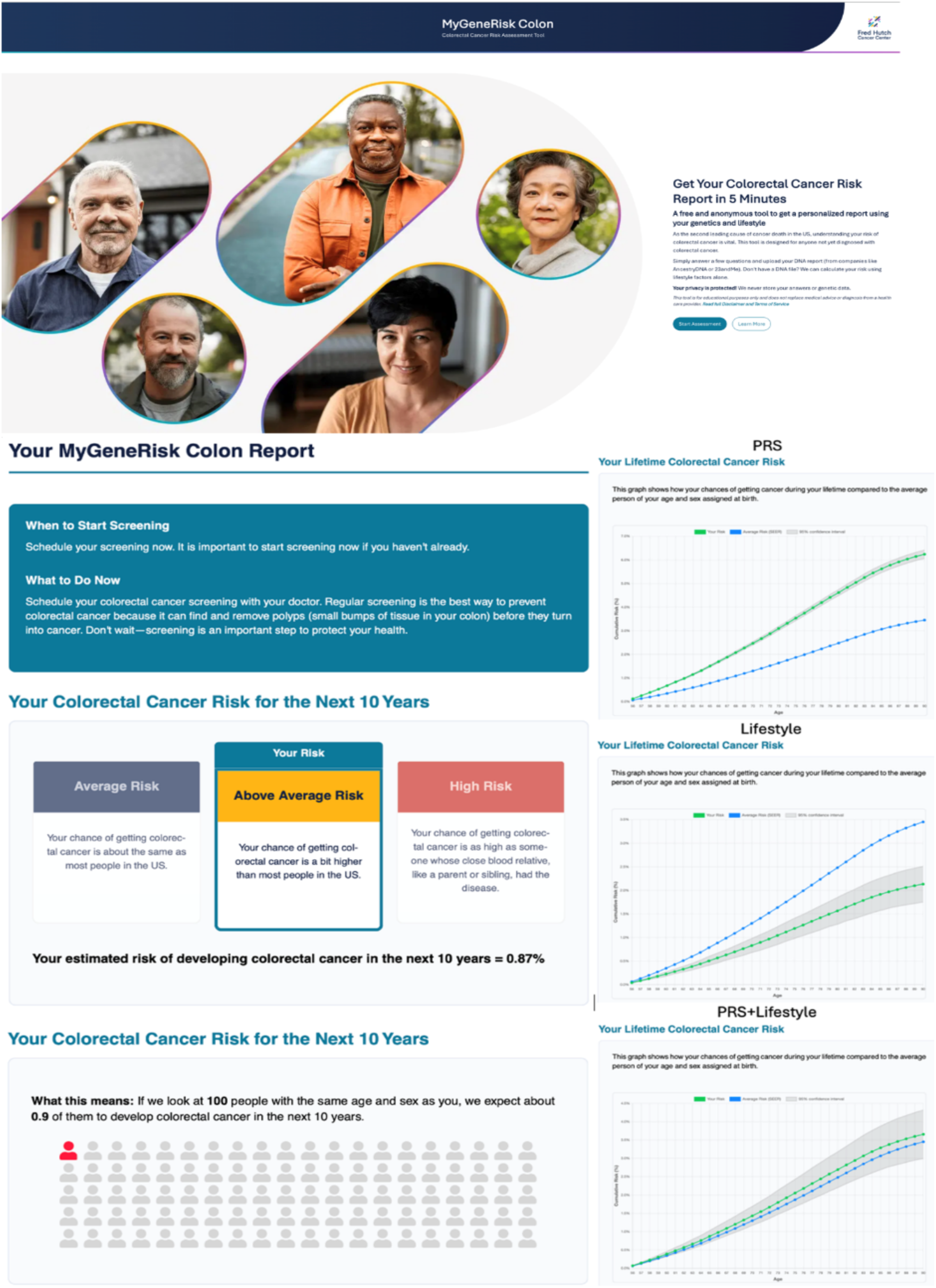
The website of the CRC risk assessment tool MyGeneRisk Colon. The top panel displays the landing page, the bottom left panel shows the first page of a sample report, and the bottom right panel presents lifetime risk portions of three sample reports using PRS only (top), lifestyle factors only (middle), and both PRS and lifestyle factors (bottom). The web tool can be accessed at https://mygenerisk-colon.fredhutch.org/ and an example of a full risk report is provided in Supplemental Materials.

### Web-based Tool Development

We developed the **MyGeneRisk Colon** web-based tool (https://mygenerisk-colon.fredhutch.org/) with emphasis on three key components: a content development process guided by community engagement through a Community Advisory Panel (CAP) with members from across the US; a technical infrastructure enabling rapid and secure computation of personalized CRC risk estimates; and data security and ethical oversight to ensure privacy and regulatory compliance. Together, these efforts created a tool that is scientifically rigorous, privacy-centered, and secure.

To ensure the web-based tool aligns with community member preferences and maximizes its benefit to the community, at the outset of our study, we formed a CAP comprised of 8 members recruited from across the U.S. through outreach efforts from cancer center and hospital community engagement offices. CAP members were community leaders engaged in diverse efforts to educate their local communities about health-related topics and were well positioned to provide important community perspectives. The CAP was diverse in terms of sex, race, ethnicity, and educational background or profession. Initial sessions focused on bidirectional education opportunities and engagement, with emphasis on PRS-related topics. Thereafter, CAP members tested three publicly available CRC risk tools ([Colorectal Cancer Risk Assessment from the University of Rochester Medical Center](https://www.urmc.rochester.edu/encyclopedia/content?contenttypeid=42&contentid=ColorectalCancerRisk), [Qcancer from ClinRisk Ltd](https://www.qcancer.org/)., and [My CancerIQ: Colorectal Cancer Assessment from Cancer Care Ontario](https://www.mycanceriq.ca/Cancers/Colorectal)), completed an electronic survey providing feedback on these tools, and participated in a facilitated discussion.

The CAP members provided the following recommendations for the new tool: 1) include an introductory paragraph explaining the tool’s purpose and a disclaimer that it is not intended for symptomatic individuals, 2) structure risk factor questions as yes/no whenever feasible, 3) include race and ethnicity, 4) provide clear actionable steps, such as speaking to a doctor about cancer screening or reducing red meat intake, following receipt of a risk score, 5) ensure that the tool takes fewer than ten minutes to complete, and 6) include a progress tracker at the top of the screen to indicate completion status. CAP members also advised on strategies and approaches for communicating CRC risk and risk factors to the users.

Based on their evaluations, several recommendations were incorporated into the design of our tool (more details are provided in the Methods section). For example, with CAP input, we developed a web tool questionnaire (included in the supplement) based on survey items used in the cohort and nested case-control studies. We also designed a personalized risk report following best practices in risk communication,^30^ and incorporated lessons learned from previous qualitative research on patient understanding of PRS,^31^ as well as the use of personalized risk information,^31^ and CAP feedback. **Figure 3** illustrates the landing page of the website, along with portions of sample reports. Full reports are provided in the Supplemental Materials.

We have architected and built a robust web-based tool infrastructure that allows real-time risk calculations. To compute individualized PRS for CRC, users first upload their raw DTC-GT data, which they need to download from the DTC site. To support real-time PRS calculation, we developed a cloud-supported, parallelized pipeline that performs preprocessing, phasing, imputation, PRS calculation, and absolute risk estimation, integrating both genetic and lifestyle information to generate a personalized risk report for each user. The system was deliberately designed to maximize privacy protection, while ensuring cost efficiency and ease of maintenance by outsourcing computationally intensive calculations to Amazon Web Services (AWS) Lambda, which provides secure, on-demand computing. No user data are stored beyond ephemeral (temporary) computation, and final risk calculations occur in the user’s browser only. Data transfer between client and AWS Lambda’s ephemeral compute environment is encrypted. PRS calculations are partitioned by chromosome, and no backend component ever has access to the complete set of genotypes. Aggregation of chromosome-specific PRS results, risk estimation and report rendering occur entirely within the user’s browser. Through this architecture, we achieve high-performance computation with minimal data exposure.

The development and deployment of the MyGeneRisk Colon platform followed rigorous standards for data security and ethical compliance. The study and tool infrastructure were reviewed and approved by Fred Hutch’s Legal Counsel, including the Office of Risk Compliance and Information Security.

## Discussion

We developed MyGeneRisk Colon, a secure, web-based tool that delivers personalized CRC risk estimates directly to individuals in real-time. By integrating user-uploaded genetic data from DTC-GT services with users’ responses to a lifestyle questionnaire, the tool calculates absolute risk using rigorously developed genome-wide PRS and lifestyle risk estimates derived from large, well-characterized, and diverse epidemiological cohorts, together with population incidence rates from SEER. Its privacy-preserving design offers a proof of concept for securely delivering individualized risk to a growing population of DTC-GT users. As genomic data becomes more integrated into healthcare, this tool provides a scalable approach for clinical decision support, enabling risk-stratified screening, facilitating more informed and collaborative discussions between patients and clinicians, and supporting more personalized cancer prevention and surveillance strategies.

Efforts are already underway to evaluate the integration of genomic information into clinical settings. For example, the Electronic Medical Records and Genomics (eMERGE) Network (https://emerge-network.org/) is conducting a prospective, pragmatic trial involving ∼24,000 individuals of diverse ancestry to evaluate whether returning genome-informed risk assessments that include PRS will lead to changes in disease management and outcomes. As genomic data are increasingly integrated into electronic health records, tools like ours could support real-time delivery of individualized risk information in both research and healthcare settings. Further research is needed to better understand patient and provider perspectives, to determine whether personalized risk prediction improves screening uptake, particularly in high-risk individuals, and to evaluate the cost-effectiveness of personalized prevention strategies.

Existing PRS tools, including those for CRC, typically focus on genetic risk alone, while most public-facing^32,33^ CRC risk calculators rely solely on lifestyle factors. Given that CRC risk arises from both genetic and lifestyle factors, an integrated approach can enhance the accuracy of prediction. Our tool uniquely combines a genome-wide PRS with self-reported lifestyle and demographic information to return CRC risk estimates tailored to the individual. To improve generalizability, we have developed our lifestyle risk assessment based on diverse populations and utilized a PRS that was developed across different ancestry populations. In addition, the use of absolute risk estimates provides a more actionable interpretation of risk than percentile-based PRS, which can be challenging to contextualize clinically.

By appropriately accounting for potentially heterogeneous effects across large and diverse studies, the estimated relative risks of lifestyle factors are expected to be generalizable to the broader population. To obtain these estimates, we employed several rigorous methods. Missing data were handled using multiple imputation treating studies as clusters, and heterogeneity in effects was accommodated through individual participant data meta-analysis with random effects. We estimated CRC risk and derived both confidence intervals and prediction intervals to quantify uncertainty of the estimation and variability in effects across studies and populations.

As no analytical methods were available for this setting, we developed and implemented a novel sampling-based approach to estimating attributable risks, which were subsequently used to calculate absolute risk. Owing to heterogeneity across studies, the precision of lifestyle-based absolute risk estimates is somewhat reduced. Future work aimed at improving the measurement of lifestyle exposures and identifying novel predictors may further refine these risk models. The accuracy of risk prediction could also be enhanced through integration of high-penetrance genes and fecal immunochemical tests (FIT).

Community engagement played a central role in the development of this tool. We collaborated with a Community Advisory Panel comprising representatives from diverse geographic areas and racial and ethnic backgrounds to ensure broad relevance. The panel’s input informed the design of the questionnaire, the structure of the report, and the communication strategies. We tailored the language, visuals, and educational resources to enhance health literacy and address various preferences^34,35^, thereby promoting broader adoption of digital risk assessment tools. The collaboration enhanced the accuracy of our communication to a broader audience regarding prediction results and their associated benefits and risks, making the tool more generalizable and acceptable across diverse populations.^36,37^

Our tool also offers several novel technical features. First, it performs advanced, real-time per-sample genotype imputation, an uncommon capability among public-facing platforms, with high concordance with traditional batch-based imputation, as detailed in the Supplemental Materials. Second, it is built on a secure, serverless infrastructure using state-of-the-art cloud services such as Amazon Web Services Lambda, enabling cost-effective, on-demand computing. It enabled us to develop a sustainable and secure web tool, one that would not have been otherwise affordable for development and maintenance. This also avoids the storage of genetic or lifestyle data, mitigating privacy concerns. While we recognize that some users may still feel uncomfortable sharing data, even temporarily, we maintain transparency and provide clear information about how we handle data. Third, it has a user-friendly interface. The web portal seamlessly handles user-submitted DTC genotype files and lifestyle survey responses. Within minutes, individuals receive personalized risk estimates, graphical summaries, and prevention recommendations (e.g., diet, screening).

We acknowledge several limitations. First, certain self-reported variables, such as diet, are subject to measurement errors. While our model incorporates this uncertainty through parameter estimation and sampling, additional research is warranted to improve exposure assessment and to further elucidate how these exposures impact CRC development. Second, although the PRS has been validated in four major racial and ethnic populations, its predictive accuracy has not yet been evaluated in other groups, including some with known elevated CRC risk, such as Native American and Alaska Native populations. While the reference panel, the 1000 Genomes Project, includes diverse populations, the individuals of mixed ancestries remain underrepresented. This underrepresentation may reduce single-sample imputation accuracy. Future methodological refinements, such as aggregating genotypes into small ad hoc pseudo-cohorts may improve population representation and imputation quality without substantially increasing computational burden, while preserving the real-time performance required for PRS calculation. Similarly, although the lifestyle risk assessment was developed across four racial and ethnic groups, non-Hispanic White individuals comprise most participants across the included studies. Third, while our model integrates externally validated PRS and well-established lifestyle risk factors, further evaluation of the combined model in broader and more diverse population settings would strengthen confidence in its generalizability and real-world applicability.

In summary, we established a secure, publicly available web portal for CRC risk assessment that jointly incorporates genetic and lifestyle factors. Through meaningful community engagement, we designed a tool that is accessible and relevant to diverse populations. With tens of millions of individuals in the U.S. alone already possessing DTC-GT data, this privacy-preserving tool demonstrates the feasibility of delivering actionable, personalized risk estimates directly to users. As genomic data become increasingly available through DTC-GT and progressively integrated in electronic health records, the framework presented here provides a model for potential clinical decision support and highlights the promise of more personalized, data-informed approaches to cancer prevention and early detection in both public health and clinical care.

## Methods

The development of the MyGeneRisk Colon web tool combined three core components: rigorous statistical modeling, community-informed design, and innovative technical implementation. This Methods section details how these elements were integrated.

First, we developed a comprehensive risk prediction model. To obtain robust and generalizable estimates of lifestyle relative risk, we conducted an individual participant data meta-analysis across sixteen diverse cohorts. Missing data were addressed using multiple imputation to reduce bias and preserve statistical efficiency. We then incorporated a genome-wide PRS and developed a novel sampling-based method to estimate attributable risk and absolute risk. Second, we used a community-engaged process to guide tool development. Our Community Advisory Panel (CAP) played a central role in shaping the questionnaire and the design of personalized reports, ensuring relevance and usability. Finally, we implemented a secure, serverless architecture designed for scalability and privacy. A key technical innovation is a high-performance pipeline that performs real-time, per-sample genotype imputation, an essential capability for a public-facing platform.

The CRC risk prediction model itself is based on a Cox proportional hazards model, which comprises two components: hazard ratios (HR) for risk factors and the baseline hazard function. The first three subsections describe how we estimated HRs. The 4^th^ and 5^th^ subsections detail how we derived the baseline hazard function by estimating the attributable risk and combining it with the population CRC incidence rate from SEER. In the 6^th^ subsection, we incorporated competing risks (i.e., mortality from other causes) and constructed the final absolute risk prediction model.

### Multiple imputation

We performed multiple imputation separately by sex (male and female) and study type (cohort and nested case-control) using joint modeling. This approach allows simultaneous imputation of both continuous and discrete variables at the participant and study levels, and accommodate data that are missing systematically or sporadically.^38^ Imputation was conducted for all risk factors with missing values, except for endoscopy (colonoscopy or sigmoidoscopy) history. Because a substantial proportion of participants in multiple studies had missing information on prior endoscopy, imputing this variable could have introduced additional uncertainty. Instead, we created an ‘unknown’ category. Each participant was therefore classified as ‘yes’, ‘no’, or ‘unknown’, ensuring that all individuals retained an observed value for this variable in the analysis. We generated 180 imputed datasets using a joint modeling approach that incorporated all risk factors and outcomes. The joint model assumed the observed data followed a multivariate normal distribution, with a latent normal structure used to handle categorical variables. Each study was treated as a cluster. Several fully observed variables were included as predictors in the imputation model, including CRC status, age at study entry, endoscopy history, and race and ethnicity. For cohort studies, we also included age at diagnosis (for CRC cases) or age at last observation (for non-cases). For nested case-control studies, we included the age at diagnosis for cases and the matched age at selection for controls. For nested case-control data, we performed 2000 burn-in iterations, and then generated 180 imputed data sets, each spaced 2,000 iterations apart. Due to the substantially large sample size in cohort studies, multiple imputation requires much more computing resources. Therefore, we used 200 for both burn-in and between-imputation updates.

For continuous variables, a logarithm transformation was applied, where appropriate, before joint modeling to accommodate the normality assumption. The imputed values were then exponentiated to return them to their original scale. For variables with a minimal value of zero, such as red meat intake, a small constant (equal to 5% quantile of the non-zero original data) was added before the logarithm transformation prior to imputation. After imputation, the same constant was subtracted after the exponentiation transformation. Thus, the value after logarithmic transformation was well defined. For continuous variables, since a random draw from a normal distribution can be arbitrarily large or small, the last step was to winsorize values to the observed range of the original data.

We performed multiple imputation using the R package *jomo*.^39^

### Individual participant data meta-analysis

We conducted an individual participant data meta-analysis to estimate sex-specific effects (HRs) of the risk factors. This approach preserves the clustering of individuals within studies and is regarded as the ‘gold standard’ for systematic review.^40^ A two-step meta-analytic approach was used.^41^ **Figure 4** shows a schematic overview of the meta-analysis workflow.

**Figure 4.**
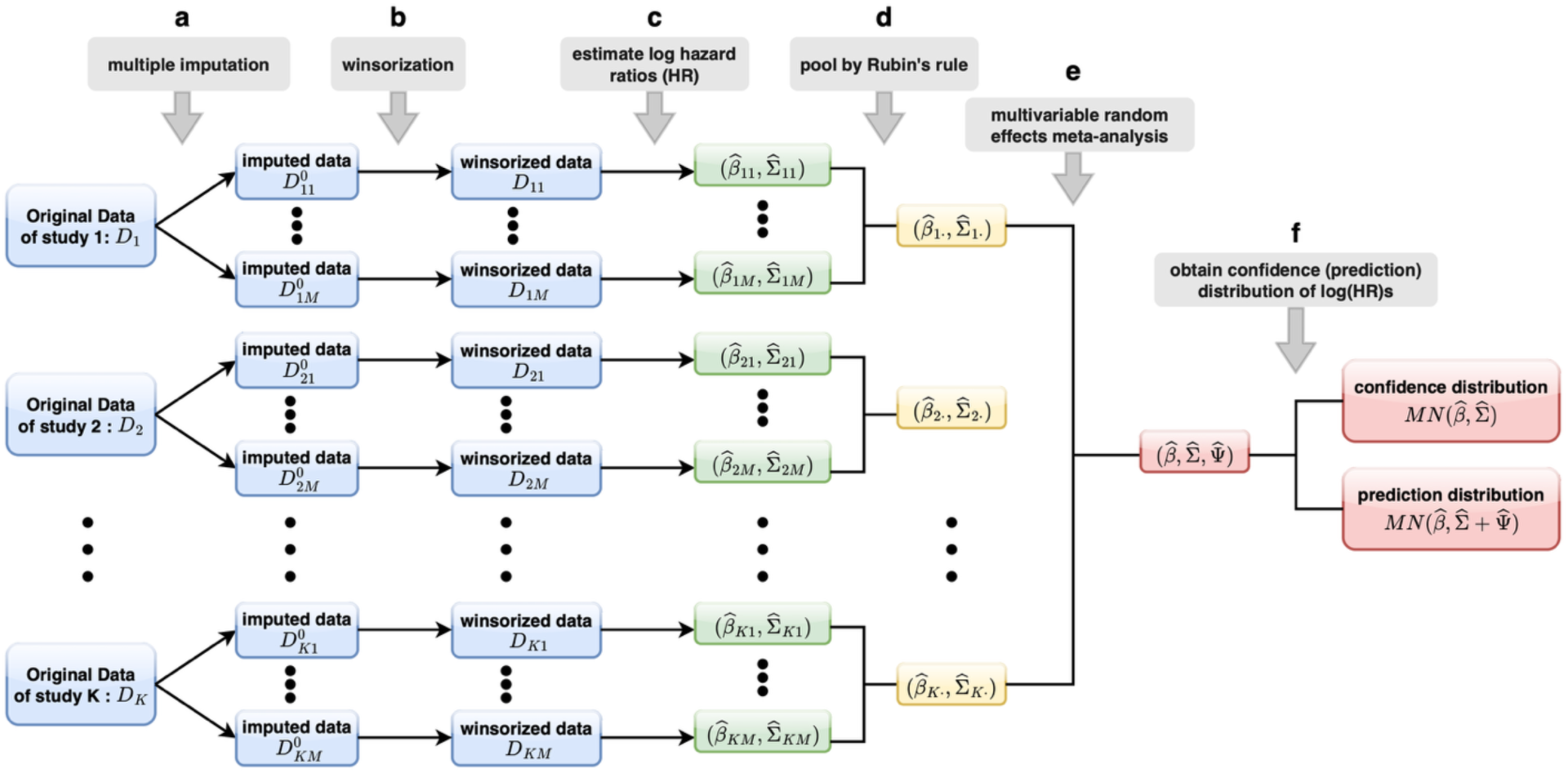
Workflow of the meta-analysis for estimating the associations of lifestyle factors with colorectal cancer. **a**. For each study, 𝑘 (𝑘 = 1, ⋯ , 𝐾), multiple imputation was performed to generate 𝑀 imputed datasets, denoted by 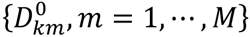. **b.** For each 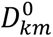, winsorization was applied as appropriate, resulting in datasets 𝐷_𝑘𝑚_. **c.** For each 𝐷_𝑘𝑚_, log(HR) estimates and (co)variance matrix were obtained by fitting a logistic (nested case-control studies) or Cox (cohort studies) regression model, including all lifestyle factors in one model, while adjusting for age. **d**. Within each study 𝑘, the estimates were pooled across multiple datasets using Rubin’s rule in a multivariate manner to account for the uncertainty of missing data imputation, yielding study-wise log(HR) estimates *β̂_k_* and (co)variance matrix *Σ̂_k_*. **e**. A multivariable random effects meta-analysis was performed to aggregate the study-specific estimates. **f**. The confidence (prediction) distribution of log(HR) estimates follows a multivariable normal distribution with mean given by the point estimate of the aggregated effects coefficients *β̂*, and (co)variance matrix *Σ̂* (and the between-study (co)variance matrix of random effects 𝛹̂). Abbreviation: log(HR), logarithm of hazard ratio.

In the first step, the imputed individual participant data were analyzed for each study. For each of the 180 imputed datasets within each study, we fit either a logistic regression model for nested case-control study or a Cox proportional hazards regression model for cohort study, including all the risk factors and adjusting for race and ethnicity as a covariate. For nested case-control studies, the logistic regression model additionally adjusted for age at study entry and time since study entry. For cohort studies, the Cox model accounted for age at study entry through left truncation, with survival outcomes defined using age as the time scale: age at diagnosis for CRC cases and age at last observation for non-cases, along with CRC status. Given the limited sample size in some studies and to enhance interpretability and ensure model stability, we included only linear effects for the risk factors. In the nested case-control analyses, we estimated odds ratio (OR), which approximates HR well for rare diseases such as CRC ^42^. For each imputed dataset, we obtained a vector of the log(HR) estimates for the risk factors and the corresponding (co)variance matrix. In some studies with limited sample size, certain risk factors were not estimable in one or more imputed datasets. To maintain consistency across imputations, if a risk factor was not estimable in any of the 180 imputed datasets, it was treated as non-estimable for that study. Using Rubin’s rule for multivariable multiple imputation, we pooled the log(HR) estimates across imputed datasets, yielding study-specific HR estimates and (co)variance matrices that accounted for the uncertainty due to missing data.^43^ Let 𝛽 be the vector of parameters of interest. Specifically, for 𝑚 = 1, … , 𝑀, 𝛽^𝑚^ denotes the estimated parameter vector from the 𝑚th imputed dataset and 𝑉𝑎𝑟(𝛽^𝑚^) denotes the corresponding (co)variance matrix. The pooled estimate of 𝛽 is simply the average 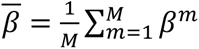 with its (co)variance matrix as

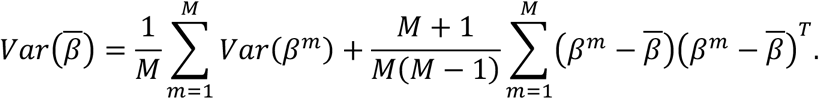

In the second step, we performed a multivariate random-effects meta-analysis to synthesize the study-specific HRs, allowing for between-study heterogeneity by incorporating random study effects.^44^ We used the restricted maximum likelihood method, assuming a diagonal between-study (co)variance structure, that is, random effects for different log(HR) parameters were assumed to be uncorrelated across studies. The method also accommodates risk factors for which HRs were not estimable in some studies. Specifically, it sets missing HR estimates to 0, corresponding covariances to 0, and variances to the largest observed variance multiplied by 10^6^, so that the corresponding estimates contribute only negligibly to the final estimates. We obtained the overall mean HR estimates and the corresponding (co)variance matrix. We derived both the confidence and prediction intervals of HR estimates. For confidence intervals, the (co)variance of the estimates is the (co)variance matrix of overall mean HR estimate reflecting its uncertainty, and for prediction intervals, it includes additionally the heterogeneity (co)variance matrix of HR estimates across studies, providing an estimate of the range of effects that might be expected in a new study setting.^45^ We performed multivariate random-effects meta-analysis using the R package *mvmeta*.^46^

### Polygenic risk score

We used the Asian-Euro PRS previously developed based on 69,175 individuals of Asian ancestry (21,731 cases; 47,444 controls) and 185,616 individuals of European ancestry (78,473 cases; 107,143 controls)^16^. The PRS was validated in independent datasets across four major self-reported groups: Asian, Black or African American, Hispanic, and Non-Hispanic White^16^. The score is constructed as a weighted sum of 1,020,293 HapMap3 SNPs, with weights derived using LDpred2^47^. These weights corresponded to the posterior mean effect sizes of SNPs, estimated from a joint Bayesian modeling of SNP effects, assuming a prior mixture distribution consisting of a point mass at zero and a normal distribution for non-zero effects. The model was trained using marginal association summary statistics from a combined Asian and European GWAS using the inverse variance estimator. It incorporated a linkage disequilibrium matrix constructed from both Asian and European individuals, weighted proportionally to their respective contributions to the combined summary statistics. A schematic overview is shown in **Figure 5**.

**Figure 5.**
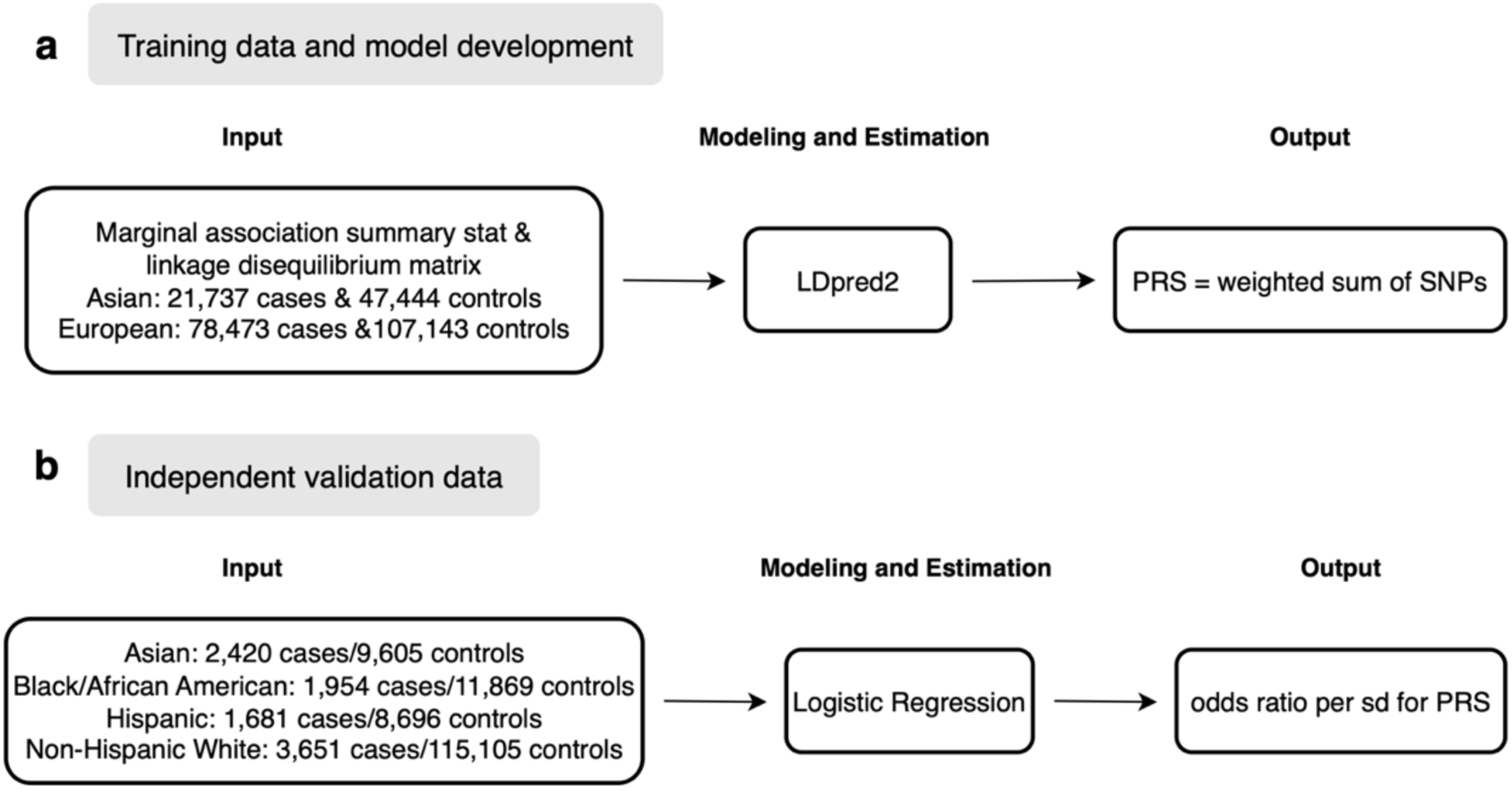
Schema for obtaining the odds ratios for PRS. (a) Training data and model development; (b) Independent validation data.

As our study populations used to estimate the effects of lifestyle factors were part of the training dataset used to develop the Asian-Euro PRS, re-estimating PRS effects using these data would lead to overfitting and potential inflated estimates. To avoid this bias, we used odds ratio estimates for the PRS obtained from independent datasets as reported in Thomas et al. (2023, **Table 3**), stratified by self-reported population groups. All analyses in Thomas et al. were adjusted for age and the top four principal components to account for potential population substructure.

### Single-sample genotype imputation

^48^Because genotype imputation in most settings is performed jointly across many samples to leverage shared haplotypes, it can be challenging to adapt these methods for per-user calculations in a web-based environment. Since PRS values are computed immediately upon upload, we implemented a single-sample imputation workflow that can process individual uploads quickly and independently. We used the well-characterized, publicly available 1000 Genomes Phase 3 as reference panel, which includes 661 African, 347 Admixed American, 504 East Asian, 503 European, and 489 South Asian. ^48^ To evaluate the performance of this single-sample genotype imputation approach, we analyzed 830 custom genotyped samples from the Personal Genome Project^49^, including 6 African, 265 Admixed American, 36 East Asian, 523 European, and 25 South Asian samples. Each sample was imputed twice: first using the single-sample workflow as implemented in the MyGeneRisk pipeline, and second using a batch workflow in which all 830 samples were jointly phased with Eagle2 and imputed with Minimac4 against the same 1000 Genomes reference panel. PRS was calculated from the dosage files from both pipelines. PRS estimates derived from single-sample and batch-imputed genotypes showed high concordance (Pearson correlation > 0.98; **Figure S4**), indicating that the single-sample workflow provide results comparable to standard batch-based methods.

### Attributable risk (AR) estimation

To develop an absolute risk model, we need to estimate attributable risks for both PRS and lifestyle factors such that the baseline hazard can be calculated.

### Deriving the ARs of lifestyle risk factors

To facilitate the estimation of ARs for lifestyle risk factors, we define a *lifestyle risk score* (LRS), constructed as a weighted sum of lifestyle risk factors with weight equal to the logarithm of their corresponding HR. For some studies, due to limited sample sizes, it was not possible to obtain HR estimates for certain lifestyle risk factors, making it impossible to estimate study-specific attributable risks (ARs) based solely on their data. To address this limitation, we developed an alternative sampling-based approach, as depicted in **Figure 6**. Using this framework, we derive both 95% confidence intervals and prediction intervals for the AR estimates as follows.

**Figure 6.**
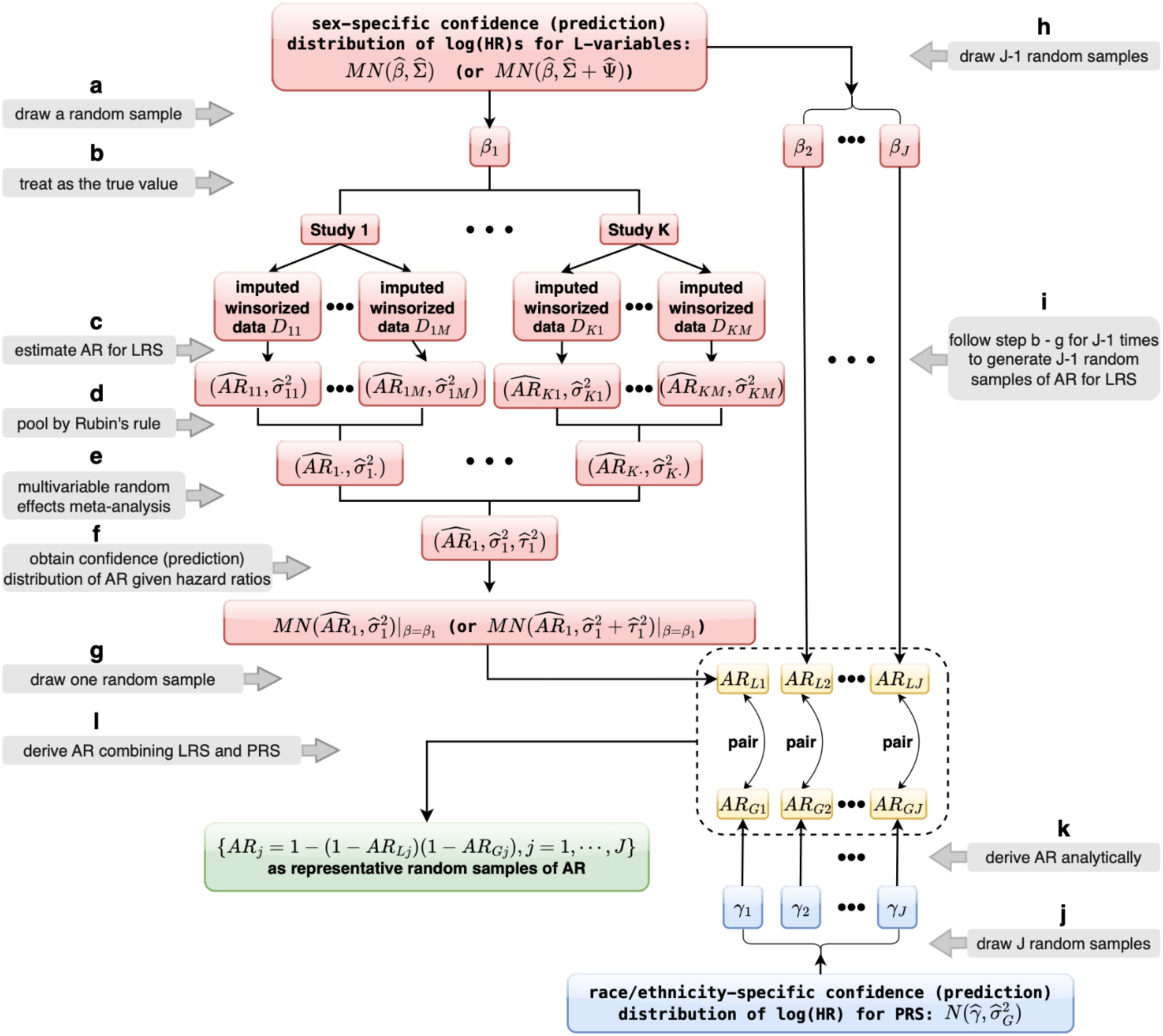
Workflow for the sampling-based estimation of attributable risks (ARs) for lifestyle risk score (LRS) and polygenic risk score (PRS). *Deriving AR for LRS*: **a**. We generated a random vector of sex-specific log(HR) values for all lifestyle risk factors from the confidence (prediction) distribution of the log(HR) estimates, denoted 𝛽_1_. These represent plausible log(HR) parameter values for predicting CRC risk in a new individual. **b-d**. Given 𝛽_1_ (treated as the truth), we obtained the study-wise confidence distribution of AR based on the multiple imputed data sets, in a manner similar to HR estimation (see Figure 4). **e-f**. For each sex and race/ethnic group, we obtained the confidence (prediction) distributions of ARs given 𝛽_1_. **g**. We randomly drew a value from the confidence (prediction) distribution of AR, denoted by 𝐴𝑅_𝐿1_ , as the AR estimate corresponding to 𝛽_1_ . **h-i**. Repeating this process for 𝐽 − 1 additional draws, {𝛽_j_, 𝑗 = 2, ⋯ , 𝐽} yielded 𝐽 pairs of LRS-specific log(HR) vector and AR estimate, {(𝛽_j_, 𝐴𝑅_𝐿j_), 𝑗 = 1, ⋯ , 𝐽}. *Deriving the ARs of PRS*: **j**. To account for the uncertainty in the PRS log(HR) estimates, we generated 𝐽 random samples from their racial/ethnic specific confidence (prediction) distribution, {𝛾_j_, 𝑗 = 1, ⋯ , 𝐽}. **k**. For each 𝛾_j_ (treated as the truth), we analytically calculated 𝐴𝑅_𝐺j_ = 1 − ∫ exp(−𝛾_j_𝑥) 𝜙(𝑥)𝑑𝑥, where 𝜙(𝑥) is the normal density function with mean 𝛾_j_ and variance 1, yielding 𝐽 pairs of PRS-specific log(HR) and AR estimates, {(𝛾_j_, 𝐴𝑅_𝐺j_), 𝑗 = 1, ⋯ , 𝐽}. *Deriving the overall ARs*: **l**. the overall ARs was calculated as {𝐴𝑅_j_ = 1 − (1 − 𝐴𝑅_𝐿j_)(1 − 𝐴𝑅_𝐺j_), 𝑗 = 1, ⋯ , 𝐽}. Abbreviations: log(HR), logarithm of hazard ratio; L-variables, lifestyle risk factors.

First, we generated 200 random samples of sex-specific log(HR) values for lifestyle risk factors, drawn from the confidence distribution of HR estimates, denoted by {𝛽_j_, 𝑗 = 1, ⋯ ,200}. Each sample, as a vector, represents a plausible set of log(HR) values for a new individual (in this case, the user of the webtool). With each sampled log(HR) vector treated as the truth, we obtained the confidence distribution of AR based on multiple imputed data sets generated using the same approach as in HR estimation (**Figure 4**). We obtained the AR confidence distributions by sex for each of racial and ethnic groups, Asian, Black or African American, Hispanic, Non-Hispanic White, as well as for the overall population. Finally, we randomly drew an AR value from its confidence distribution, as a plausible AR value corresponding to the specific log(HR) vector. Overall, we yielded 200 LRS-specific AR estimates, each linked to one of the 200 LRS-specific log({(𝛽_j_, 𝐴𝑅_𝐿j_), 𝑗 = 1, ⋯ ,200}.

For prediction distribution of AR, we followed the same procedure except that the sex-specific log(HR) values would be drawn from the prediction distribution of HR estimates, and we then obtained the prediction distribution of AR.

*Deriving the ARs of PRS*: To account for uncertainty in the log(HR) estimates of PRS for Asian, Black or African American, Hispanic, and non-Hispanic White, respectively, we generated 200 random draws from the population-specific distribution of PRS-specific log(HR) estimate, denoted by {𝛾_j_, 𝑗 = 1, ⋯ ,200}. Based on Thomas et al. (2023)^16^, the distribution of PRS among cases followed a normal distribution and was observed to be homogenous across studies. For a given HR value for PRS, the PRS-specific AR can therefore by derived analytically by *AR_G_* = 1 − ∫ exp(− log(𝐻𝑅) 𝑥) 𝜙(𝑥)𝑑𝑥, where 𝜙(𝑥) is the normal density function of PRS among cases with mean log(𝐻𝑅) and variance 1. We generated 200 PRS-specific AR estimates, each linked to one of the 200 sampled log(HR) values, yielding paired estimates {(𝛾_j_, 𝐴𝑅_𝐺j_), 𝑗 = 1, ⋯ ,200}.

### Deriving the overall ARs of PRS and lifestyle factors (LRS)

In our study populations, PRS and LRS were only weakly correlated (Pearson correlation coefficient < 0.1), indicating that genetic and lifestyle factors predict CRC risk independently. Therefore, we calculated the overall AR as a function of the LRS-specific AR and the PRS-specific AR, 𝐴𝑅 = (1 − 𝐴𝑅_𝐿_)(1 − 𝐴𝑅_𝐺_).

Specifically, for each combination of sex and race and ethnicity, we generated 200 random draws of paired PRS-specific AR and LRS-specific AR, i.e., {(𝐴𝑅_𝐿j_, 𝐴𝑅_𝐺j_), 𝑗 = 1, ⋯ ,200}, with each pair linked to one of the afore-mentioned LRS-specific log(HR) vector and PRS-specific log(HR) {(𝛽_j_, 𝛾_j_), 𝑗 = 1, ⋯ ,200}. We calculated the overall AR by {𝐴𝑅_j_ = 1 −(1 − 𝐴𝑅_𝐿j_)(1 − 𝐴𝑅_𝐺j_), 𝑗 = 1, ⋯ ,200} and obtained 200 triples {(𝛽_j_, 𝛾_j_, 𝐴𝑅_j_), 𝑗 = 1, ⋯ ,200} that are used to calculate absolute risk. This will be done for both the calculation of confidence intervals and prediction intervals.

### Absolute risk estimation

To estimate an individual’s future risk of developing CRC, we used sex- and race and ethnicity-specific HR and sex-, age-, and race and ethnicity-specific baseline hazard function estimates based on the individual’s PRS and lifestyle risk profile. Accounting for competing risks, we calculated the probability of developing CRC over a prespecified time interval (e.g., 5, 10, or 20 years), conditional on current age and risk factors.

Specifically, we computed sex-, age-, and race and ethnicity-specific baseline hazard rates by multiplying the corresponding Surveillance Epidemiology and End Results (SEER) CRC incidence rates averaged over 2015 to 2019 by 1-AR (as described above). For individuals who prefer not to answer their race and ethnicity, we also calculated sex-specific baseline hazard rate by using the sex-specific SEER rates with sex-specific AR. This procedure was repeated for each of the 200 sampled log(HR) parameter sets and their corresponding AR estimates, i.e., {(𝛽_j_, 𝛾_j_, 𝐴𝑅_j_), 𝑗 = 1, ⋯ ,200}. For each set, we calculated the absolute risk estimates based on the individual’s profile. From these 200 absolute risk values, we calculated the mean as the final risk estimate with 95% confidence or prediction intervals calculated using the standard error under a normal approximation.

The formula for calculating the absolute risk is described as follows.^50^ Let 𝑡_0_ be the individual’s current age, (𝛽, 𝛾) represent the log(HR) estimates for lifestyle risk factors and PRS, and (𝐿, 𝐺) denote the individual’s risk profile with Lifestyle factors 𝐿 and PRS 𝐺, respectively. Let 𝜆_0_(𝑢) be the baseline hazard rate of CRC at age 𝑢, and 𝜆_𝑜𝑐_(𝑢) be the SEER mortality rate for causes other than CRC at age 𝑢. The absolute risk of developing CRC within 𝑡 years for a disease-free individual aged 𝑡_0_ is given by

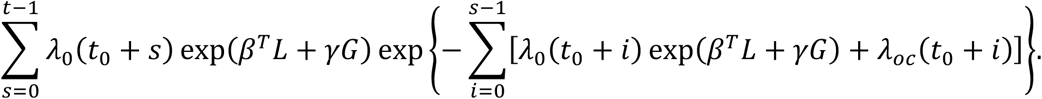

Taken together, we provided the following risk estimates:

• **Individual risk estimates**

Individual risk was calculated using sex-, race-, and ethnicity-specific SEER CRC rates, sex-specific HR estimates for lifestyle factors (**Table 2**), and race- and ethnicity-specific HR estimates for PRS (**Table 3**). If an individual selected “Choose not to answer” for sex, we calculated risk estimates for both males and females and reported both. If an individual selected “Choose not to answer” for race and ethnicity, we used overall SEER rates across all racial and ethnic groups by sex, and for PRS we used the HR for all groups (**Table 3**). We then reported the corresponding absolute risk based on the individual’s reported sex, or both male and female estimates if sex was also not provided.

• **Average risk estimates**

For comparison with the general population, we computed absolute risk using overall SEER CRC rates averaged from 2015 -2019. The rates were obtained by combining data across males and females and across all racial and ethnic groups. We provide this as the average population risk by age.

### Engagement of Community Advisory Panel

To ensure our research aligns with community member preferences and maximizes its benefit to the community, we formed a Community Advisory Panel (CAP) with members recruited from across the U.S. at the beginning of the project. This initiative was supported by the Community Outreach and Engagement (COE) leaders from four collaborating institutions: Fred Hutchinson Cancer Center (Washington), Moffitt Cancer Center (Florida), Cedars-Sinai Medical Center (California), and Cleveland Clinic (Ohio). We collaborated closely with each institution’s COE team to recruit community members directly or to present the project to standing institutional community advisory boards. Details of the recruitment process and meetings are provided in the Supplementary Materials. In brief, CAP meetings offer bidirectional educational opportunities and engagement, where CAP members engage in facilitated discussions with the CAP research leads. To prepare the CAP members for collaboration in the research project, the CAP research leads organized training on CRC, CRC screening, precision medicine, PRS, and cost-effectiveness to optimize screening. These interactive sessions established the foundation to equip the CAP to provide recommendations on the development of this risk prediction tool. The primary role of the CAP was to advise on webtool development, focusing on the overall approach, questionnaire, and report, drawing on their expertise in working with their communities.

### Development of questionnaire and personalized risk reports

We developed a questionnaire based on survey items used in the cohort and nested case-control studies ^19^ with input from CAP members. The questionnaire included all variables described in the Lifestyle Factors section and added symptom-related questions to help identify users who may need to consult a healthcare provider instead of using the webtool. We provided detailed explanations on meat consumption, endoscopic procedures, and over-the-counter supplements to enhance understanding for users. Users may choose not to answer questions about sex, or race and ethnicity. If sex is not provided, CRC risk estimates for both male and female are displayed.

If race and ethnicity are missing, the tool presents sex-specific CRC risk estimates for the overall population. In this scenario, we used sex-specific SEER overall incidence rates along with the following parameter estimates: sex-specific HRs and corresponding ARs for lifestyle factors (with ARs pooled across racial and ethnic groups); and PRS ORs and corresponding analytically-derived AR that were not stratified by sex or race and ethnicity.

We designed a personalized risk report following best practices in risk communication,^30^ and incorporated lessons learned from previous qualitative research on patient understanding of PRS,^31^ and use of personalized risk information,^31^ as well as CAP feedback. The CAP feedback on the report included input on 1) use of visual graphics to depict CRC risk, 2) descriptions of CRC screening options, 3) importance of colors, font size, and layout to be compliant with American Disabilities Act guidance, 4) ability to receive portable document format (PDF) of personalized report, and 5) call to action to speak with a doctor about their results. The report included graphs that depicted both individual and average 10-year and lifetime risks of CRC. We also provided educational information on managing CRC risk through actionable steps, such as initiating CRC screening, increasing physical activity and consuming fruits and fiber, or reducing tobacco use and red meat consumption. Prototypes of the personalized report were improved based on CAP input. We collaborated with patient education experts to further enhance the clarity and effectiveness of our messaging for the broader population. A sample report is available in the Supplemental Materials.

### Web tool infrastructure

We developed a cloud-supported, parallelized pipeline to rapidly compute individualized PRS for CRC from DTC-GT data and combined these with lifestyle risk estimates. The pipeline performs preprocessing, phasing, imputation, PRS calculation, and absolute risk estimation, integrating both genetic and lifestyle information to generate a personalized risk assessment for each user. The tool does not store data other than during computation, and final scores are calculated in the user’s browser, ensuring no data or results are stored for longer than necessary. Users can choose to only upload DTC-GT data to assess their genetic risk, only answer the questionnaire to assess their lifestyle risk, or do both for the most comprehensive risk assessment. The time from uploading the DTC-GT data to providing the raw PRS ranges from 2 to 3 minutes, depending on upload speed. Users can answer the lifestyle questions while the PRS is calculated in the cloud. Completing the lifestyle questionnaire typically takes between 3 and 6 minutes. The calculations of the adjusted PRS and/or lifestyle risk, as well as the report generation, take place in the user’s browser in seconds.

Upon uploading the DTC-GT data by the user, the input file is first validated to ensure it is in a supported format and genotyping data are of sufficient quality. This includes checks for structural integrity (e.g., required columns and valid SNP identifiers), verification that the number of SNPs per chromosome meets or exceeds expected counts, reducing the risk of truncated or corrupted files. Files that fail this check are rejected without further processed, and the user is notified with an explanation. Once validated, the raw DTC-GT file is parsed by chromosome. Only autosomal variants (chromosomes 1–22) are retained, because PRS is calculated only from these loci. Variants on sex chromosome and mitochondrial DNA are excluded. Each autosomal chromosome is then processed independently using a cloud-native function, enabling parallel execution. Details on the QC checks are provided in the Supplemental Materials.

Specifically, we utilize Amazon Web Services (AWS) Lambda, an on-demand serverless compute service optimized for ephemeral workloads. Ephemeral storage is provided as a temporary directory that exists only during the Lambda function’s execution. This storage is automatically deleted once the function ends, ensuring that no data is retained after processing. To not solely rely on Amazon’s deletion protocol, the Lambda function deletes data as soon as it’s no longer needed. AWS Lambda was selected not only for its cost-effective model, but also for its robust security profile: each Lambda function executes in a stateless, short-lived sandbox environment with no access to any operating system files or storage from other functions, thereby substantially reducing the risk of data leakage or unauthorized access. Furthermore, each Lambda instance processes a single chromosome and calculates only a partial, chromosome-specific PRS value.

Within each Lambda instance, genotype data are converted from the vendor-specific format to the widely used Variant Call Format (VCF). Variants with missing genotypes or structural variation are filtered out. The tool automatically determines the genome build of the uploaded genotyping file and will lift it over to the build used for phasing and imputation, if necessary. Cleaned VCFs are phased using Eagle2^51^ (version 2.4.1), which infers haplotype structure based on local linkage disequilibrium patterns. The phased variants are then imputed using Minimac4^52^ (version 4.1.3) using the 1000 Genomes reference panel, allowing untyped variants to be inferred with probabilistic confidence.

Following imputation, each Lambda function calculates a chromosome-specific raw PRS by summing the imputed dosages weighted by their corresponding effect sizes. These per-chromosome PRS values are temporarily stored until all 22 scores are ready to be sent to the browser, where they are summed to generate the overall raw PRS, which is calibrated based on genetic ancestry using a reference distribution derived from the 1000 Genomes panel.^16^

The calibrated PRS score is then used to compute the personalized estimate of 10-year and lifetime CRC risks, along with lifestyle and demographic factors from the questionnaire. The risk calculations are performed locally in the user’s browser. At no point are questionnaire responses or final risk estimates transmitted or stored remotely. A personalized report is generated for the user, including estimated 10-year and lifetime CRC risks up to the age of 90, along with a comparison to population averages. The report also provides general guidance on CRC screening options and risk mitigation strategies.

### Data Security and Ethical Compliance

The development and deployment of our web-based prediction tool adhered to rigorous standards for data security and ethical compliance. To ensure data security, we used industry-standard encryption protocols for file transfer, decentralized and ephemeral compute and storage solutions, and access controls aligned with the principle of least privilege. The tool is hosted on AWS and managed by a dedicated team at Fred Hutch. The AWS platform undergoes continuous monitoring by Fred Hutch and third-party organizations and receives updates to security protocols to ensure the ongoing protection of data. Importantly, data are deleted as soon as PRS calculation is finished or within 10 minutes if an error occurred; no individual-level data is retained, linked to other personal information, or stored for longer than necessary for report generation. Prior to its release, our webtool was reviewed internally to ensure compliance with medical device requirements. Before uploading data and answering questions, all users must acknowledge that they have reviewed the disclaimer stating the site is not intended to provide medical advice and indicate their acceptance of the terms and conditions regarding privacy. Our study was approved by Fred Hutchinson’s IRB, and a full description of the approval can be found in the supplemental files.

## Data availability

In accordance with the informed consent and IRB approval, all data are made available via controlled access. Individual-level genetic and demographic data from all studies except UK Biobank are deposited in dbGaP (accession nos. phs001415.v1.p1, phs001315, v1.p1, phs001078.v1.p1, phs001903.v1.p1, phs001856.v1.p1 and phs001045.v1.p1). UK Biobank genotype, demographic, and lifestyle data are available through http://www.ukbiobank.ac.uk. Access to individual-level lifestyle data is controlled through oversight committees of each study and is coordinated through the Genetics and Epidemiology of Colorectal Cancer Consortium (GECCO, contact), which will respond to a data request within two weeks. From that point in time, it is expected that approvals will take up to two months.

## Acknowledgements

We thank Claire Betz, Victoria Biyani, Laura Carr, Theary Chhim, Natalie Curtis, Jennifer Davies, Blake C Douglass, Eleanor Englund, Jeremy Fisher, Antonio Leon, Sam Minot, Shawntá Mosley-App, Adina Mueller, Patrick Shelby, John Soltys, John Wenning, Kim Westphal, Kat Wynn, and Michael Zager, for helpful input and for their careful reading and suggestions on early versions of the manuscript.

## **(a)** Funding Support

Genetics and Epidemiology of Colorectal Cancer Consortium (GECCO): National Cancer Institute, National Institutes of Health, U.S. Department of Health and Human Services R01 CA244588, P50 CA285275, R01 CA273198, R01 CA 297681. Genotyping/Sequencing services were provided by the Center for Inherited Disease Research (CIDR) contract number HHSN268201700006I and HHSN268201200008I. This research was funded in part through the NIH/NCI Cancer Center Support Grant P30 CA015704 and NIH/NHGRI HG008657. Scientific Computing Infrastructure at Fred Hutch funded by ORIP grant S10OD028685. This work was supported (in part) by the Thomas B. and Jeannette E. Laws McCabe Fund at the University of Pennsylvania to Jiayin Zheng.

BWHS: National Institutes of Health U01 CA164974. The authors would like to acknowledge the contribution to this study from central cancer registries supported through the Centers for Disease Control and Prevention’s National Program of Cancer Registries (NPCR) and/or the National Cancer Institute’s Surveillance, Epidemiology, and End Results (SEER) Program. Central registries may also be supported by state agencies, universities, and cancer centers. Participating central cancer registries include any of the following: Alabama, Alaska, Arizona, Arkansas, California, Colorado, Connecticut, Delaware, Florida, Georgia, Hawaii, Idaho, Indiana, Illinois, Iowa, Kentucky, Louisiana, Massachusetts, Maine, Maryland, Michigan, Mississippi, Montana, Nebraska, Nevada, New Hampshire, New Jersey, New Mexico, New York, North Carolina, North Dakota, Ohio, Oklahoma, Oregon, Pennsylvania, Rhode Island,, South Carolina, Tennessee, Texas, Utah, Virginia, West Virginia, Wyoming. The authors assume full responsibility for analyses and interpretation of these data.

CLUE II funding was from the National Cancer Institute (U01 CA086308, Early Detection Research Network; P30 CA006973), National Institute on Aging (U01 AG018033), and the American Institute for Cancer Research

CPS-II: The American Cancer Society funds the creation, maintenance, and updating of the Cancer Prevention Study-II (CPS-II) cohort.

Harvard cohorts: HPFS is supported by the National Institutes of Health (P01 CA055075, UM1 CA167552, U01 CA167552, R01 CA137178, R01 CA151993, and R35 CA197735), NHS by the National Institutes of Health (P01 CA087969, UM1 CA186107, R01 CA137178, R01 CA151993, and R35 CA197735), and PHS by the National Institutes of Health (R01 CA042182). Effort of S. Ogino related to the GECCO and Harvard cohorts was additionally supported by the American Cancer Society Clinical Research Professor Award (CRP-24-1185864-01-PROF).

Japan Public Health Center-based Prospective Study was supported by the National Cancer Center Research and Development Fund (23-A-31[toku], 26-A-2, 29-A-4, 2020-J-4, and 2023-J-4) (since 2011) and a Grant-in-Aid for Cancer Research from the Ministry of Health, Labour and Welfare of Japan (from 1989 to 2010).

MCCS: Melbourne Collaborative Cohort Study (MCCS): MCCS cohort recruitment was funded by VicHealth and Cancer Council Victoria. The MCCS was further supported by Australian National Health and Medical Research Council grants 209057, 396414 and 1074383 and by infrastructure provided by Cancer Council Victoria. Cases and their vital status were ascertained through the Victorian Cancer Registry (VCR).

MEC: National Institutes of Health (R37 CA054281, P01 CA033619, R01CA126895, and U01 CA164973).

PLCO: Intramural Research Program of the Division of Cancer Epidemiology and Genetics and supported by contracts from the Division of Cancer Prevention, National Cancer Institute, NIH, DHHS. Funding was provided by National Institutes of Health (NIH), Genes, Environment and Health Initiative (GEI) Z01 CP 010200, NIH U01 HG004446, and NIH GEI U01 HG 004438.

SCCS: National Institutes of Health U01 CA202979

SMC_COSM (Swedish Mammography Cohort and Cohort of Swedish Men): This work is supported by the Swedish Research Council / SIMPLER Research Infrastructure grant, the Swedish Cancer Foundation, and the Karolinska Institute’s Distinguished Professor Award to Alicja Wolk.

UK Biobank: This research has been conducted using the UK Biobank Resource under Application Number 8614

VITAL: National Institutes of Health (K05 CA154337).

WHI: The WHI program is funded by the National Heart, Lung, and Blood Institute, National Institutes of Health, U.S. Department of Health and Human Services through contracts 75N92021D00001,75N92021D00002, 75N92021D00003, 75N92021D00004, 75N92021D00005

## **(b)** Study participants and data contributions

CLUE II: We thank the participants of Clue I and Clue II and appreciate the continued efforts of the staff at the Johns Hopkins George W. Comstock Center for Public Health Research and Prevention in the conduct of the Clue Cohort Studies.

Maryland Cancer Registry (MCR): Cancer data were provided by the Maryland Cancer Registry, Center for Cancer Prevention and Control, Maryland Department of Health, with funding from the State of Maryland and the Maryland Cigarette Restitution Fund. The collection and availability of cancer registry data is also supported by the Cooperative Agreement NU58DP007114, funded by the Centers for Disease Control and Prevention. Its contents are solely the responsibility of the authors and do not necessarily represent the official views of the Centers for Disease Control and Prevention or the Department of Health and Human Services.

CPS-II: The authors express sincere appreciation to all Cancer Prevention Study-II participants, and to each member of the study and biospecimen management group. The authors would like to acknowledge the contribution to this study from central cancer registries supported through the Centers for Disease Control and Prevention’s National Program of Cancer Registries and cancer registries supported by the National Cancer Institute’s Surveillance Epidemiology and End Results Program. The study protocol was approved by the institutional review boards of Emory University, and those of participating registries as required. The authors assume full responsibility for all analyses and interpretation of results. The views expressed here are those of the authors and do not necessarily represent the American Cancer Society or the American Cancer Society – Cancer Action Network.

Harvard cohorts: The study protocol was approved by the institutional review boards of the Brigham and Women’s Hospital and Harvard T.H. Chan School of Public Health, and those of participating registries as required. We acknowledge Channing Division of Network Medicine, Department of Medicine, Brigham and Women’s Hospital as home of the NHS. The authors would like to acknowledge the contribution to this study from central cancer registries supported through the Centers for Disease Control and Prevention’s National Program of Cancer Registries (NPCR) and/or the National Cancer Institute’s Surveillance, Epidemiology, and End Results (SEER) Program. Central registries may also be supported by state agencies, universities, and cancer centers. Participating central cancer registries include the following: Alabama, Alaska, Arizona, Arkansas, California, Colorado, Connecticut, Delaware, Florida, Georgia, Hawaii, Idaho, Indiana, Iowa, Kentucky, Louisiana, Massachusetts, Maine, Maryland, Michigan, Mississippi, Montana, Nebraska, Nevada, New Hampshire, New Jersey, New Mexico, New York, North Carolina, North Dakota, Ohio, Oklahoma, Oregon, Pennsylvania, Puerto Rico, Rhode Island, Seattle SEER Registry, South Carolina, Tennessee, Texas, Utah, Virginia, West Virginia, Wyoming. The authors assume full responsibility for analyses and interpretation of these data.

PLCO: The authors thank the PLCO Cancer Screening Trial screening center investigators and the staff from Information Management Services Inc and Westat Inc. Most importantly, we thank the study participants for their contributions that made this study possible. Cancer incidence data have been provided by the District of Columbia Cancer Registry, Georgia Cancer Registry, Hawaii Cancer Registry, Minnesota Cancer Surveillance System, Missouri Cancer Registry, Nevada Central Cancer Registry, Pennsylvania Cancer Registry, Texas Cancer Registry, Virginia Cancer Registry, and Wisconsin Cancer Reporting System. All are supported in part by funds from the Center for Disease Control and Prevention, National Program for Central Registries, local states or by the National Cancer Institute, Surveillance, Epidemiology, and End Results program. The results reported here and the conclusions derived are the sole responsibility of the authors.

SCCS: SCCS protocols were approved by Vanderbilt University Medical Center, and all participants provided written informed consent. All research was conducted in accordance with the Declaration of Helsinki and Declaration of Istanbul. Patients and the public were not directly involved in the planning, design, or conduct of the present study or SCCS, although the intake questionnaires were designed for and tailored to the SCCS population

WHI: The authors thank the WHI investigators and staff for their dedication, and the study participants for making the program possible. A full listing of WHI investigators can be found at: https://s3-us-west-2.amazonaws.com/www-whi-org/wp-content/uploads/WHI-Investigator-Long-List.pdf

This manuscript is the result of funding in whole or in part by the National Institutes of Health (NIH). It is subject to the NIH Public Access Policy. Through acceptance of this federal funding, NIH has been given a right to make this manuscript publicly available in PubMed Central upon the Official Date of Publication, as defined by NIH.

## Author contributions

Conceptualization and design: J.Z., M.T., B.A., C.G., V.S., S.T.V., G.P.J., R.L.M., I.L.V., U.P., and L.H. Data curation: J.Z., C.Q., A.T.C., J.R.H., M.I., A.W., T.Y., A.N.B.-H., W.Z., and U.P. Formal analysis: J.Z., R.S.S., and C.Q. Methodology: J.Z., B.A., J.R.H., S.T.V., A.N.B.-H., S.L.S., U.P., and L.H. Recruitment: A.T.C., S.J.C., C.A.H., L.L.M., S.C.L., S.O., C.Y.U., A.W., J.C.F., W.Z., R.J.M., and U.P. Supervision: U.P. and L.H. Writing – original draft: J.Z., R.S.S., H.Y., S.S.T., S.R.G., U.P., and L.H. Writing - review& editing: J.Z., R.S.S., H.Y., C.Q., M.T., S.S.T., C.A., B.A., D.C.C., J.K.L., S.I.B., A.T.C., S.J.C., C.G., S.R.G., C.A.H., J.R.H., M.I., L.L.M., S.C.L., J.M., I.M., S.O., V.S., C.Y.U., K.V., L.L.W., A.W., W.W., A.W., T.Y., S.T.V., G.P.J., A.N.B.-H., R.L.M., E.A.P., J.C.F., W.Z., R.J.M., J.R.P., S.L.S., I.L.V., U.P., and L.H. All authors read and approved the final manuscript.

## Supplemental Materials

- Supplemental tables and figures

- Table S1a. Descriptive characteristics of lifestyle risk factors by colorectal cancer (CRC) disease status and sex
- Table S1b. Missing counts and proportions of lifestyle risk factors by colorectal cancer (CRC) disease status and sex
- Table S1c. Winsorization cutoffs of continuous lifestyle risk factors by sex
- Figure S1: Forest plots of hazard ratios for lifestyle factors (female)
- Figure S2: Forest plots of hazard ratios for lifestyle factors (male)
- Figure S3. Scatterplot of polygenic risk scores (PRS) from per-sample imputation (X) and cohort imputation (Y)
- Harmonization of lifestyle factors
- Description of study population
- Description of CAP members and regular meetings
- Description of security and approval management
- Questionaries
- Sample report
- Pipeline QC

