## Supplemental Materials for "MyGeneRisk Colon: A Web-Based Tool for Personalized Colorectal Cancer Risk Prediction Based on Genetics and Lifestyle"

### Table of Contents


Supplemental Tables and Figures 2

Harmonization of Lifestyle Factors 11

Study Descriptions 13

Development and Description of Community Advisory Panel 17

Data Security and Approval 18

Questionnaires 19

Sample Reports 23

- PRS and lifestyle combined 26
- PRS only 32
- Lifestyle only 36

Polygenic Risk Score Pipeline and QC Checks 40

**Table S1a. Descriptive characteristics of lifestyle risk factors by colorectal cancer (CRC) disease status and sex.**

| | Female | | Male | |
|---|---|---|---|---|
| | Cases (N=9371) | Controls (N=353128) | Cases (N=7984) | Controls (N=302782) |
| **Race/ethnicity** | | | | |
| Asian | 537 (5.7%) | 8853 (2.5%) | 559 (7.0%) | 9648 (3.2%) |
| Black or African American | 574 (6.1%) | 6772 (1.9%) | 306 (3.8%) | 5225 (1.7%) |
| Hispanic | 27 (0.3%) | 1315 (0.4%) | 39 (0.5%) | 1702 (0.6%) |
| Non-Hispanic White | 8185 (87.3%) | 327454 (92.7%) | 6893 (86.3%) | 279805 (92.4%) |
| The others or missing | 48 (0.5%) | 8734 (2.5%) | 187 (2.3%) | 6402 (2.1%) |
| **Age at study entry** | | | | |
| Mean (SD) | 63.3 (8.4) | 57.8 (8.0) | 63.7 (7.9) | 58.2 (8.1) |
| **Height, cm** | | | | |
| Mean (SD) | 162.6 (6.9) | 162.6 (6.4) | 176.5 (7.6) | 176.1 (7.0) |
| **BMI, kg/m$^2$** | | | | |
| Mean (SD) | 27.3 (5.6) | 27.1 (5.3) | 27.2 (4.1) | 27.7 (4.2) |
| **Family history of CRC** | | | | |
| Yes | 1299 (15.3%) | 37756 (10.7%) | 956 (14.0%) | 31674 (10.5%) |
| **History of colonoscopy/sigmoidoscopy** | | | | |
| No | 4796 (51.2%) | 232433 (65.8%) | 4142 (51.9%) | 184143 (60.8%) |
| Unknown | 1309 (14.0%) | 5826 (1.6%) | 1423 (17.8%) | 8420 (2.8%) |
| Yes | 3266 (34.9%) | 114869 (32.5%) | 2419 (30.3%) | 110219 (36.4%) |
| **Diabetes** | | | | |
| Yes | 690 (7.6%) | 14759 (4.2%) | 736 (9.4%) | 21161 (7.0%) |
| **Smoking status, ever smoker** | | | | |
| Yes | 4342 (47.6%) | 144862 (41.3%) | 5192 (65.9%) | 162876 (54.1%) |
| **Smoking, pack-years** | | | | |
| Mean (SD) | 10.7 (18.6) | 7.2 (14.4) | 18.9 (23.7) | 13.3 (21.1) |
| **Alcohol consumption, g/day** | | | | |
| Mean (SD) | 6.1 (11.0) | 8.2 (11.0) | 17.7 (24.7) | 20.0 (23.7) |

| | | | | |
|---|---|---|---|---|
| **Aspirin use** | | | | |
| Yes | 1977 (24.1%) | 53011 (15.3%) | 2290 (35.1%) | 74019 (25.0%) |
| **NSAIDs use** | | | | |
| Yes | 1316 (16.5%) | 63320 (18.3%) | 656 (10.4%) | 37259 (12.6%) |
| **Post-menopausal hormone use** | | | | |
| Yes | 2835 (32.5%) | 56518 (16.2%) | 0 (0.0%) | 0 (0.0%) |
| **Calcium** | | | | |
| Yes | 3254 (42.4%) | 117993 (35.4%) | 604 (10.7%) | 9520 (3.4%) |
| **Processed meat, servings/day** | | | | |
| Mean (SD) | 0.2 (0.3) | 0.2 (0.2) | 0.3 (0.4) | 0.3 (0.2) |
| **Red meat, servings/day** | | | | |
| Mean (SD) | 0.7 (0.6) | 0.4 (0.3) | 0.9 (0.8) | 0.5 (0.6) |
| **Fruit, servings/day** | | | | |
| Mean (SD) | 2.3 (1.9) | 2.7 (1.8) | 2.1 (1.9) | 2.3 (1.8) |
| **Vegetable, servings/day** | | | | |
| Mean (SD) | 2.7 (2.3) | 1.3 (1.6) | 2.7 (2.4) | 1.3 (1.8) |

For categorical variables, count and percentage were calculated. For continuous variables, mean and standard deviation were calculated.

**Table S1b. Missing counts and proportions of lifestyle risk factors by colorectal cancer (CRC) disease status and sex.**

| | Female | | Male | |
|---|---|---|---|---|
| | Cases (N=9371) | Controls (N=353128) | Cases (N=7984) | Controls (N=302782) |
| **Height, cm** | | | | |
| Missing | 50 (0.5%) | 1660 (0.5%) | 83 (1.0%) | 2109 (0.7%) |
| **BMI, kg/m²** | | | | |
| Missing | 187 (2.0%) | 2677 (0.8%) | 213 (2.7%) | 2914 (1.0%) |
| **Family history of CRC** | | | | |
| Missing | 901 (9.6%) | 1422 (0.4%) | 1159 (14.5%) | 1913 (0.6%) |
| **History of colonoscopy/sigmoidoscopy** | | | | |
| Unknown | 1309 (14.0%) | 5826 (1.6%) | 1423 (17.8%) | 8420 (2.8%) |
| **Diabetes** | | | | |
| Missing | 232 (2.5%) | 1990 (0.6%) | 160 (2.0%) | 1843 (0.6%) |
| **Smoking status, ever smoker** | | | | |
| Missing | 253 (2.7%) | 1965 (0.6%) | 105 (1.3%) | 1505 (0.5%) |
| **Smoking, pack-years** | | | | |
| Missing | 622 (6.6%) | 42046 (11.9%) | 830 (10.4%) | 43897 (14.5%) |
| **Alcohol consumption, g/day** | | | | |
| Missing | 851 (9.1%) | 17862 (5.1%) | 737 (9.2%) | 19017 (6.3%) |
| **Aspirin use** | | | | |
| Missing | 1156 (12.3%) | 6948 (2.0%) | 1453 (18.2%) | 6873 (2.3%) |
| **NSAIDs use** | | | | |
| Missing | 1418 (15.1%) | 7814 (2.2%) | 1677 (21.0%) | 7413 (2.4%) |
| **Post-menopausal hormone use** | | | | |
| Missing | 642 (6.9%) | 5220 (1.5%) | - | - |
| **Calcium** | | | | |

| | | | | |
|---|---|---|---|---|
| Missing | 1698 (18.1%) | 19954 (5.7%) | 2315 (29.0%) | 21534 (7.1%) |
| **Processed meat, servings/day** | | | | |
| Missing | 589 (6.3%) | 16727 (4.7%) | 745 (9.3%) | 18184 (6.0%) |
| **Red meat, servings/day** | | | | |
| Missing | 586 (6.3%) | 16073 (4.6%) | 550 (6.9%) | 17396 (5.7%) |
| **Fruit, servings/day** | | | | |
| Missing | 765 (8.2%) | 19355 (5.5%) | 584 (7.3%) | 22082 (7.3%) |
| **Vegetable, servings/day** | | | | |
| Missing | 773 (8.2%) | 19435 (5.5%) | 579 (7.3%) | 22509 (7.4%) |

**Table S1c. Winsorization cutpoints of continuous lifestyle risk factors by sex.**

| | Female | | Male | |
|---|---|---|---|---|
| | Lower cutpoint | upper cutpoint | lower cutpoint | upper cutpoint |
| Height, cm | 150 | 175 | 162 | 191 |
| BMI, kg/m$^2$ | 19.5 | 40.4 | 20.8 | 38.0 |
| Alcohol consumption, g/day | - | 29 | - | 63 |
| Smoking, pack-years | - | 37 | - | 56 |
| Red meat, servings/day | - | 1 | - | 1.6 |
| Fruit, servings/day | - | 5.8 | - | 5.5 |

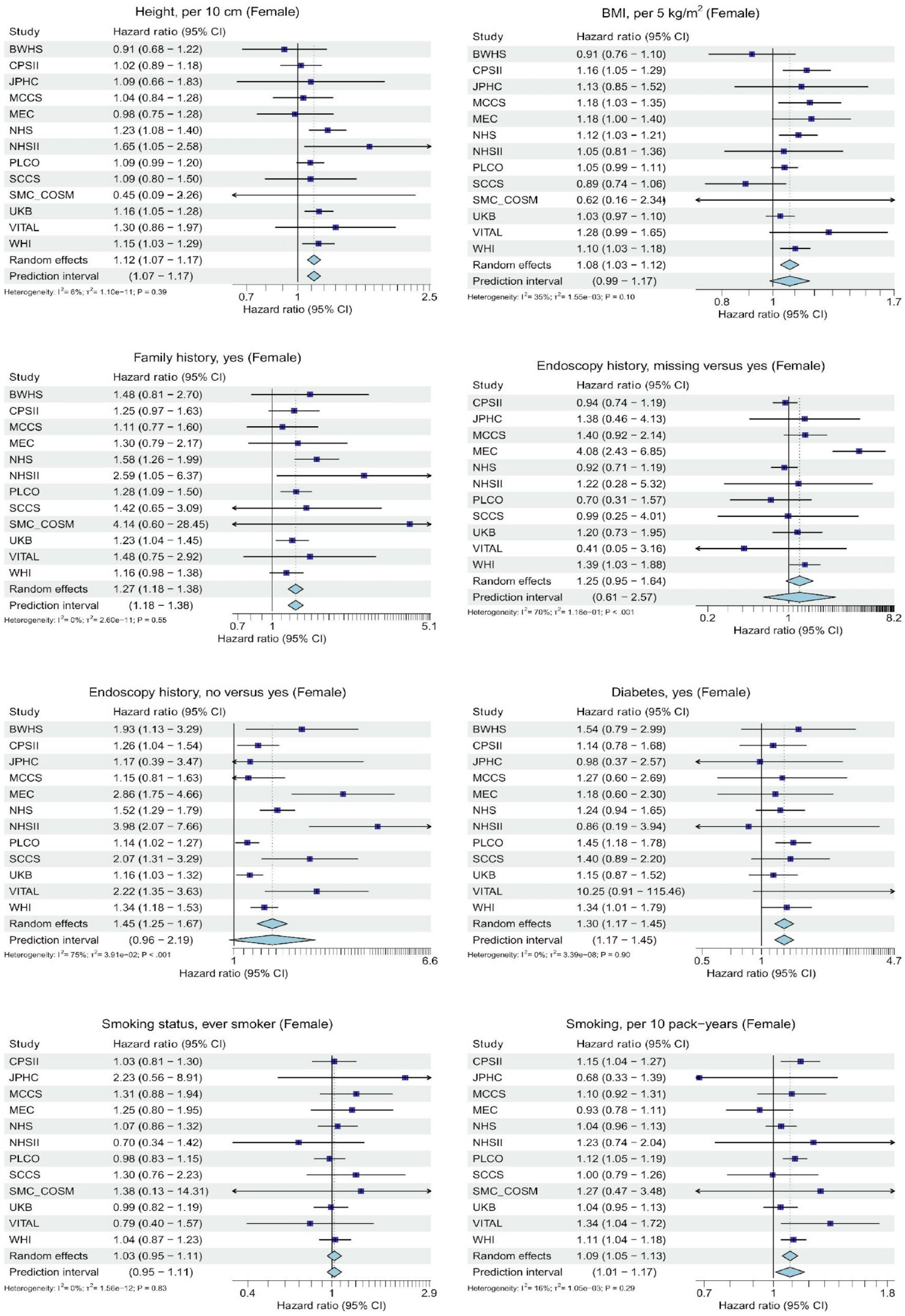


**Figure S1: Forest plots of hazard ratios for lifestyle factors (female).** In some studies, specific hazard ratios in some studies may not be estimable due to perfect separation or low event counts in certain covariate categories. CI: confidence interval.

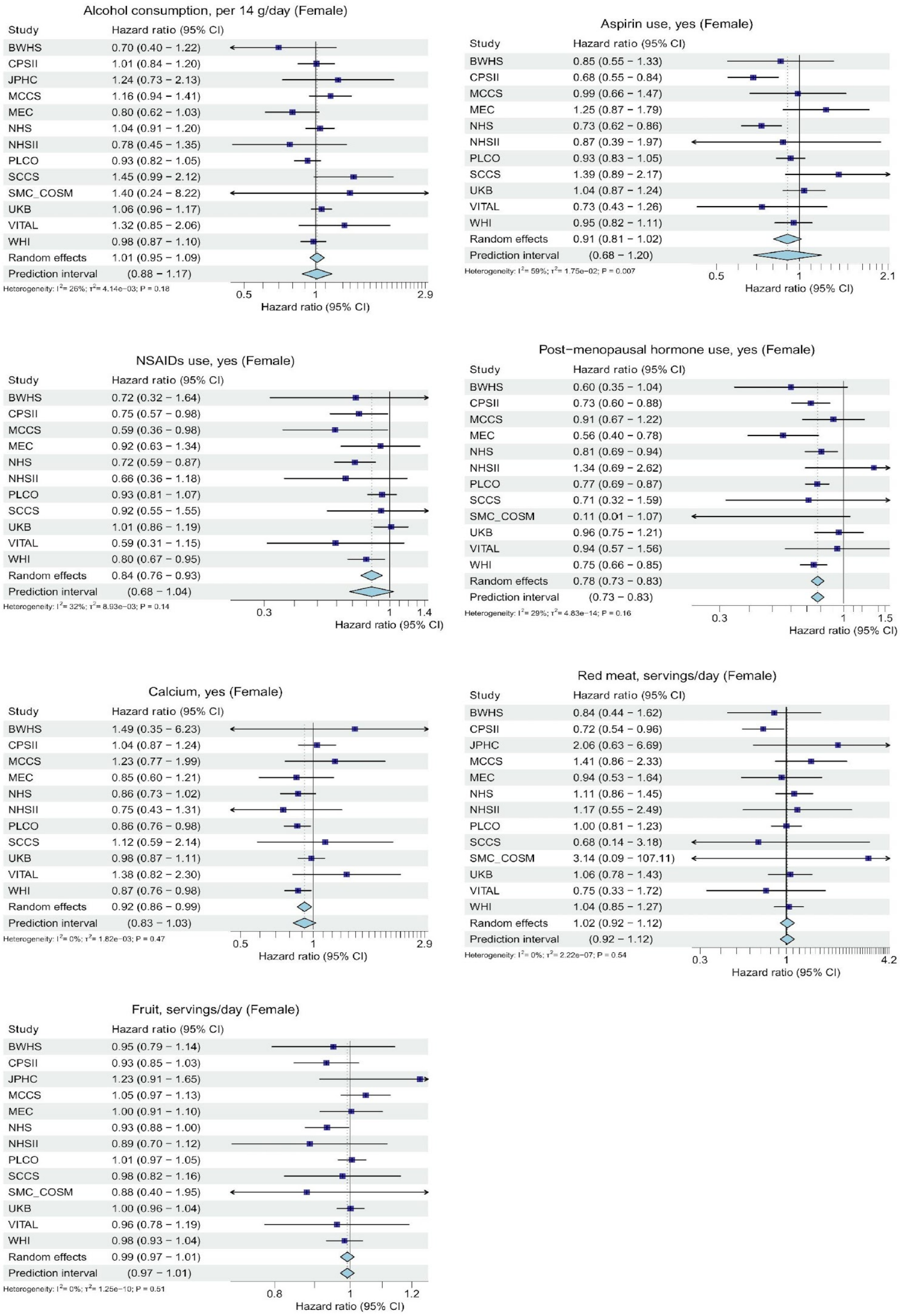


**Figure S1: Forest plots of hazard ratios for lifestyle factors (female) (continued).** In some studies, specific hazard ratios in some studies may not be estimable due to perfect separation or low event counts in certain covariate categories. CI: confidence interval.

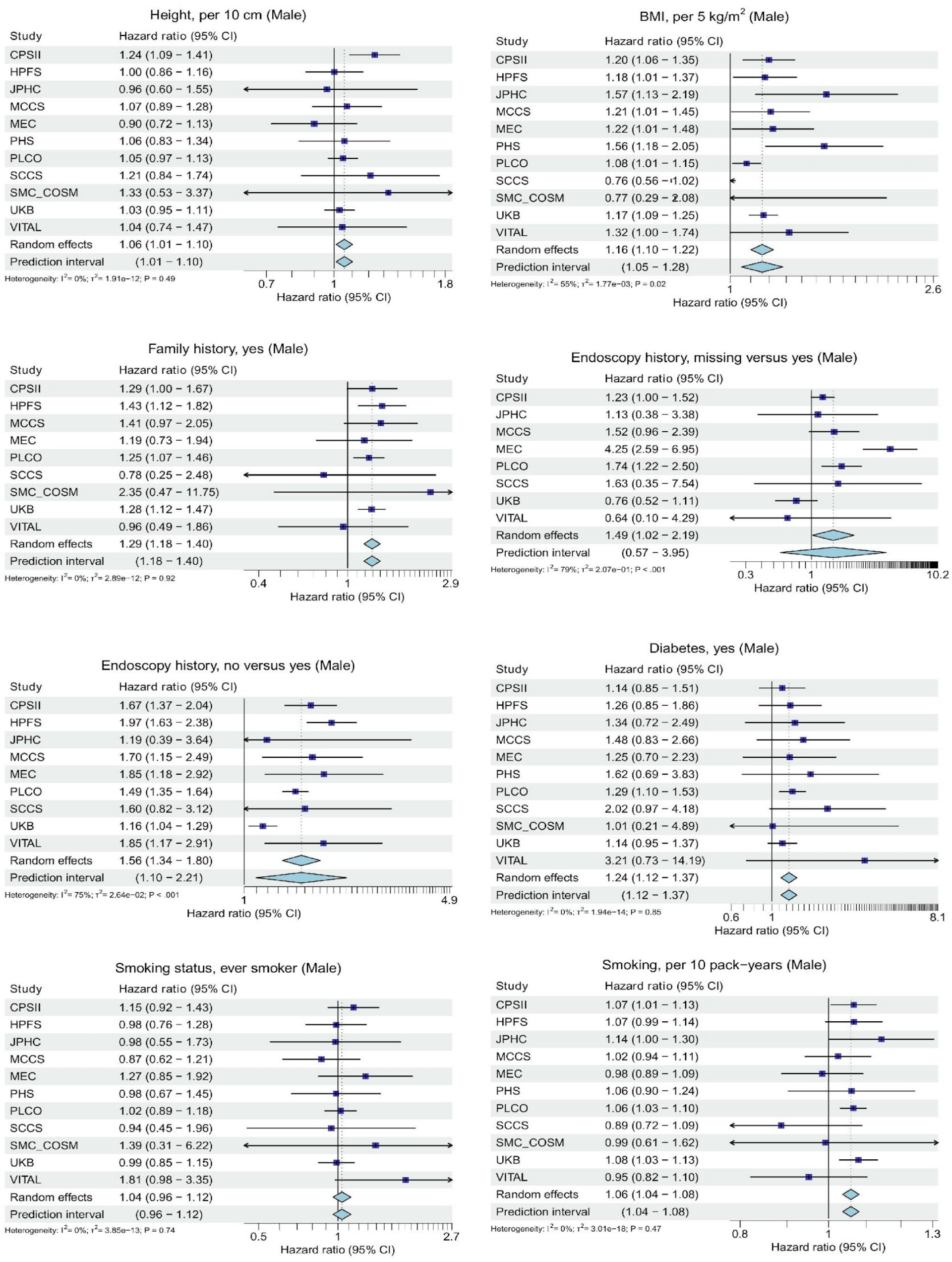


**Figure S2: Forest plots of hazard ratios for lifestyle factors (male).** In some studies, specific hazard ratios in some studies may not be estimable due to perfect separation or low event counts in certain covariate categories. CI: confidence interval.

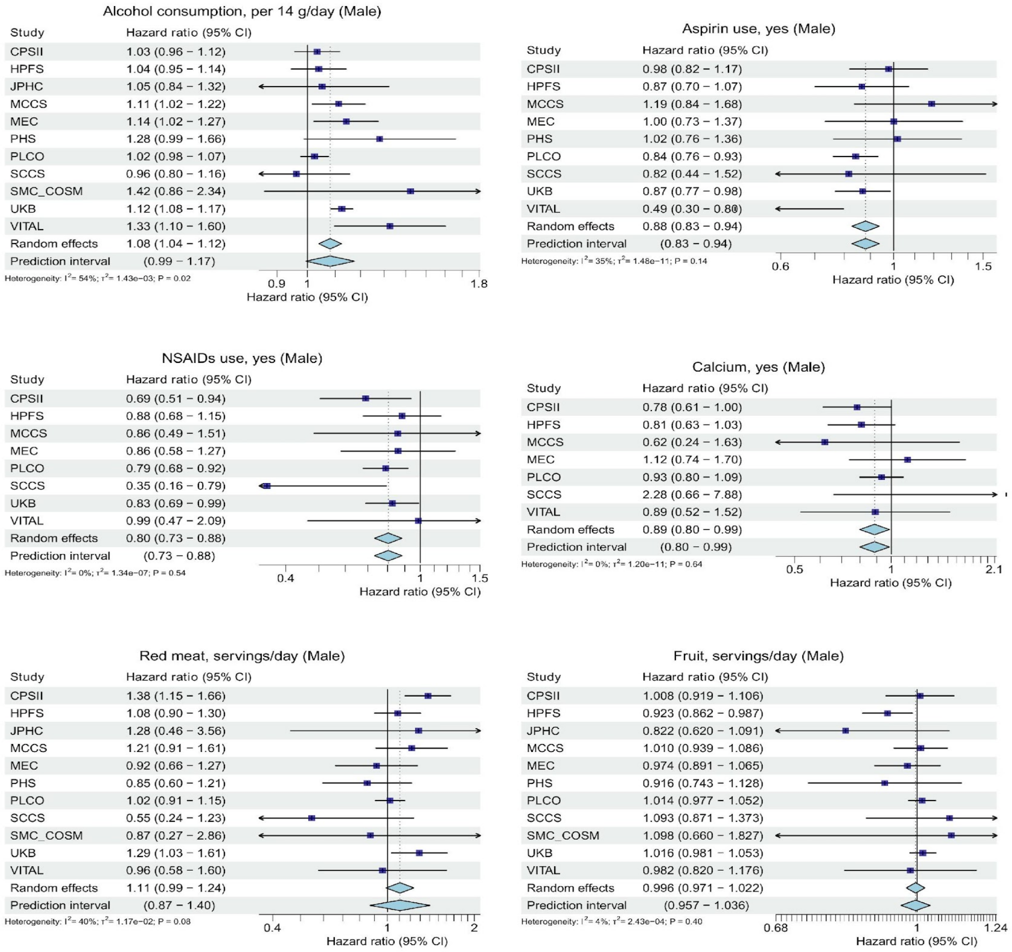


**Figure S2: Forest plots of hazard ratios for lifestyle factors (male) (continued).** In some studies, specific hazard ratios in some studies may not be estimable due to perfect separation or low event counts in certain covariate categories. CI: confidence interval.

#### Harmonization of Lifestyle Factors

To ensure all the variables are comparable across studies, individual data from all studies were harmonized at the coordinating center at the Fred Hutchinson Cancer Center using a standardized protocol (Supplementary Figure S1 in Jeon et.al., 2018).

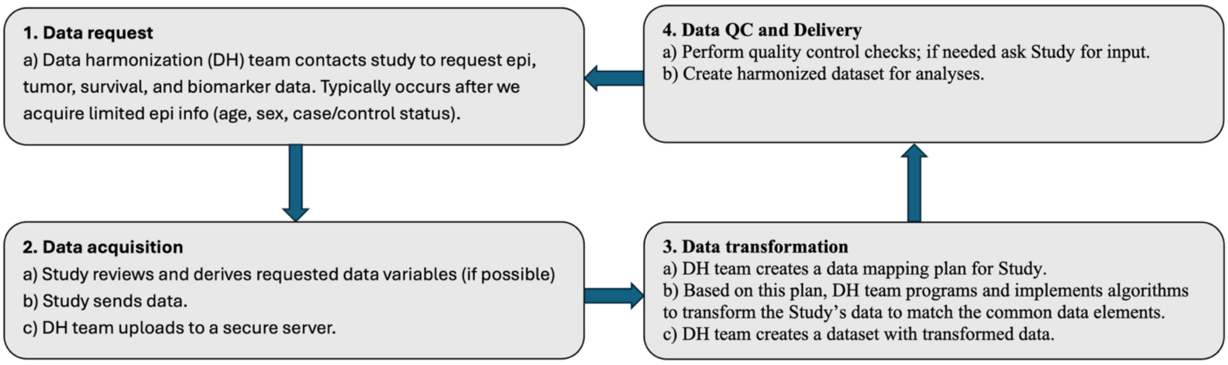


Figure S4 Overview of the data harmonization process.

Information on basic demographics and lifestyle risk factors was collected by self-report using in-person interviews and/or structured questionnaires. We carried out a multistep data harmonization procedure, reconciling each study's unique protocols and data-collection instruments (Figure S4). First, we defined common data elements. We examined the questionnaires and data dictionaries for each study to identify study-specific data elements that could be mapped to common data elements. Through an iterative process, we communicated with each data contributor to obtain relevant data and coding information. The data elements were written to a common data platform, transformed, and combined into a single dataset with common definitions, standardized permissible values, and standardized coding. The mapping and resulting data were reviewed for quality assurance, and range and logic checks were performed to assess data and data distributions within and between studies. Outlying samples were truncated to the minimum or maximum value of the established range for each variable. All variables were collected at the study reference time, which was defined as study entry or blood collection. Age at referent time was defined in years and modeled continuously.

#### Height and Body mass index

Height was defined in centimeters. Body mass index was calculated as body weight (kg) divided by height ($m^2$).

#### Family history

Family history was a yes/no variable for presence or absence of a first-degree relative with CRC.

#### Endoscopy history

Endoscopy history was coded as yes, no, or missing, depending on whether a participant had sigmoidoscopy or colonoscopy screening before the study reference time, or such information was missing.

#### Diabetes

Type 2 diabetes was coded as yes or no.

**Smoking**

Smoking status was defined as never and ever smoking; it was defined as “yes” for current or former smokers and “no” for never smokers. Pack-years of smoking were calculated by multiplying the average number of packs of cigarettes smoked per day by smoking duration (years). Smoking pack-years among ever smokers was harmonized across studies by sex- and study-specific quartiles with quartile cutoffs determined within the controls of each study and sex. For never smokers, it was assigned as “0.” This variable was treated as a continuous variable in the analysis.

**Alcohol consumption**

We converted consumption of alcoholic beverages into grams of alcohol per day by summing the alcohol content of all beverages consumed daily and treated this variable as continuous in the analysis.

**Aspirin and nonsteroidal anti-inflammatory drug use**

We used dichotomous variables for regular use of aspirin and regular use of nonsteroidal anti-inflammatory drugs (yes or no). Aspirin use is defined as “yes” if a person used aspirin regularly in the reference time period and “no” otherwise. Nonsteroidal anti-inflammatory drug use is defined as “yes” if a person used non-aspirin nonsteroidal anti-inflammatory drug regularly in the reference time period and “no” otherwise.

**Post-menopausal hormone use**

Menopausal hormone therapy use was defined as “yes” if any menopausal hormone therapy use at reference time and “no” otherwise.

**Dietary variables**

Intake of dietary factors was assessed using food frequency questionnaires. The dietary variables of fruits, vegetables, and red or processed meats were measured in servings/day; calcium supplement as mg/day. Calcium intake (mg/day) was determined from calcium in supplements (single + multivitamins + antacids) when available. For studies that entered supplement data as regular user vs nonuser, we assumed regular use was 500 mg/d, 500 mg/single tablet, or 130 mg/multivitamin tablet (the generic dose in supplements). We used a dichotomous variable for calcium supplement with “yes” for those greater than 214 mg/day and “no” for those below 214 mg/day. The cutoff of 214 mg/day for calcium supplements comes from 1500 mg/week. All dietary variables were treated as continuous variables in the analysis.

**Study Descriptions**

The following cohorts that have been harmonized as part of the Genetics and Epidemiology of Colorectal Cancer Consortium (GECCO) and are described in more detail below: Black Women's Health Study (BWHS), Campaign Against Cancer and Heart Disease (CLUEII), Cancer Prevention Study II (CPS II), Health Professionals Follow-up Study (HPFS), Japan Public Health Center-based prospective Study (JPHC), Melbourne Collaborative Cohort Study (MCCS), Multiethnic Cohort Study (MEC), Nurses' Health Study (NHS), Nurses' Health Study II (NHSII), Physician's Health Study (PHS), Prostate, Lung, Colorectal and Ovarian Cancer Screening Trail (PLCO), Southern Community Cohort Study (SCCS), Swedish Mammography Cohort and Swedish Men Cohort (SMC_COSM), UK Biobank, VITamins And Lifestyle (VITAL), and Women's Health Initiative (WHI).

**Black Women's Health Study (BWHS)**

The BWHS is a large cohort focused on the health of African American women. The purpose is to identify and evaluate causes and preventives of cancers and other serious illnesses in African American women. Among the diseases being studied are breast cancer, colorectal cancer, type 2 diabetes, uterine fibroids, systemic lupus erythematosus, and cardiovascular disease. The study began in 1995, when 59,000 black women from all parts of the United States enrolled through postal questionnaires. The women provided demographic and health data on the 1995 baseline questionnaire, including information on weight, height, smoking, drinking, contraceptive use, use of other selected medications, illnesses, reproductive history, physical activity, diet, use of health care, and other factors. The participants are followed through biennial questionnaires to determine the occurrence of cancers and other illnesses and to update information on risk factors. Self-reports of cancer are confirmed through medical records and state cancer registry records. Mouthwash-swish samples, as a source of DNA, were obtained from ~26,000 BWHS participants in 2002-2007.

**Campaign Against Cancer and Heart Disease (CLUE II)**

The Campaign Against Cancer and Heart Disease, is a prospective cohort designed to identify biomarkers and other factors associated with risk of cancer, heart disease, and other conditions. 32,894 participants were recruited from May through October 1989 from Washington County, Maryland and surrounding communities.

**Cancer Prevention Study II (CPS II)**

Men and women in the Cancer Prevention Study II (CPS-II) Nutrition Cohort (n = 184,194) were recruited from among the 1.2 million US adults enrolled in the CPS-II Cohort, a study of cancer mortality that was initiated in 1982. In 1992 and 1993, a detailed questionnaire was mailed to a subgroup of the baseline cohort. Respondents to this questionnaire were enrolled into the CPS-II Nutrition Cohort. Participants in the CPS-II Nutrition Cohort were followed for cancer incidence and mortality; they received additional mailed questionnaires in 1997 and every 2 years thereafter to update exposure information and to obtain self-reported cancer diagnoses. Self-reported cancers were verified through medical records or linkage with state cancer registries. Fatal cases were also identified through linkage with the National Death Index.

**Health Professionals Follow-up Study (HPFS)**

The HPFS cohort comprised 51,529 men aged 40-75 who, in 1986, responded to a mailed questionnaire. Participants provided information on health-related exposures, including current and past smoking history, age, weight, height, diet, physical activity, aspirin use, and family history of colorectal cancer. Colorectal cancer and the outcomes were reported by participants or next-of-kin and were followed up through review of the medical and pathology record by physicians. Overall, more than 97% of self-reported colorectal cancers were confirmed by medical record review. Information was abstracted on histology and primary location. Follow-up evaluation has been excellent, with 94% of the men responding to date.

**Japan Public Health Center-based prospective Study (JPHC)**

The Japan Public Health Center-based prospective Study conducted a baseline survey for 140,420 registered residents aged 40-69 years within 11 public health center areas nationwide in 1990-94. The questionnaire was distributed mostly by hand. Approximately 113,000 people returned the questionnaire, giving a response rate of 81%, and 48,000 provided blood samples or health checkup data, which most providing both.  Five- and 10-year follow-up surveys were conducted to update information on lifestyle habits and health conditions. In total, 130 000 participants responded to at least one of the three questionnaire surveys, and 78 000 to all three. In addition, 60 000 participants provided blood samples on at least one of the two sampling times, and 23 000 on both. The subjects have been followed for vital status and the occurrence of cancer and other diseases.

**Melbourne Collaborative Cohort Study (MCCS)**

The MCCS is a prospective study that recruited 41,514 healthy adult volunteers (17,045 men) aged between 27 and 76 years (99% aged 40-69) from the Melbourne metropolitan area between 1990 and 1994. Incident cases of invasive (including metastatic) adenocarcinoma of the colon or rectum were identified through linkage to the Victorian Cancer Registry and other State cancer registries in Australia.

**Multiethnic Cohort Study (MEC)**

MEC was initiated in 1993 to investigate the impact of dietary and environmental factors on major chronic diseases, particularly cancer, in ethnically diverse populations in Hawaiâi and California. The study recruited 96,810 men and 118,441 women aged 45 to 75 years between 1993 and 1996. Incident colorectal cancer cases occurring since January 1995, and controls were contacted for blood or saliva samples. Colorectal cancer cases are identified through the Rapid Reporting System of the Hawaiâi Tumor Registry and through quarterly linkage to the Los Angeles County Cancer Surveillance Program. Both registries are members of SEER.

**Nurses' Health Study I (NHSI)**

The Nurses' Health Study I (NHSI) cohort began in 1976 when 121,700 married female registered nurses age 30 to 55 years returned the initial questionnaire that ascertained a variety of important health-related exposures. Since 1976, follow-up questionnaires have been mailed every 2 years. Colorectal cancer and other outcomes were reported by participants or next-of-kin and followed up through review of the medical and pathology record by physicians. Overall, more than 97% of self-reported colorectal cancers were confirmed by medical-record review. Information was abstracted on histology and primary location. The rate of follow-up evaluation has been high: as a proportion of the total possible follow-up time, follow-up evaluation has been more than 92%. In 1989 to 1990, 32,826 women in NHS I mailed blood samples by overnight courier, which were aliquoted into buffy coat and stored in liquid nitrogen. In 2001 to 2004, 29,684 women in NHS I who did not previously provide a blood sample mailed a swish-and-spit sample of buccal cells.

**Nurses' Health Study II (NHSII)**

The Nurses' Health Study II (NHSII) is an ongoing cohort of 116,430 female registered nurses in the US, aged 25-42 years at baseline in 1989. Demographic, lifestyle and health-related information were obtained from participants at baseline and updated every 2 years using self-administered questionnaires. The follow-up rate in each cycle has been over 90% to date. Study participants who had not previously reported a diagnosis of cancer and had responded to the 1995 NHSII study questionnaire were invited to provide blood samples between 1996 and 1999. Blood samples were collected from 29,611 NHSII participants, aged 32 to 54 years at the time of blood draw. Similarly, between 2004 and 2006, active study participants who had not previously provided a blood sample were invited to provide buccal samples. Swish-and-spit sample of buccal cells were received from 29,859 participants. Incident cases of colorectal adenocarcinoma were ascertained first by self-report and later confirmed by reviewing medical records and pathology reports within each follow up cycle. Deaths due to colorectal cancer were identified through family or next of kin or by querying the National Death Index.

**Physician's Health Study (PHS)**

The PHS was established as a randomized, double-blind, placebo-controlled trial of aspirin and alpha-carotene among 22,071 healthy U.S. male physicians, between 40 and 84 years of age in 1982. Participants completed two mailed questionnaires before being randomly assigned, additional questionnaires at six and 12 months, and questionnaires annually thereafter. In addition, participants were sent postcards at six months to ascertain status. From August 1982 to December 1984, 14,916 baseline blood samples were collected from the physicians during the run-in phase before randomization. When participants report a diagnosis of cancer, medical records and pathology reports are reviewed by study physicians who are blinded to exposure data.

**Prostate, Lung, Colorectal and Ovarian Cancer Screening Trail (PLCO)**

PLCO enrolled 154,934 participants (men and women, aged between 55 and 74 years) at ten centers into a large, randomized, two-arm trial to determine the effectiveness of screening to reduce cancer mortality. Sequential blood samples were collected from participants assigned to the screening arm. Participation was 93% at the baseline blood draw. In the observational (control) arm, buccal cells were collected via mail using the "swish-and-spit" protocol and participation rate was 65

**Southern Community Cohort Study (SCCS)**

The SCCS is a prospective cohort investigation initiated in 2001 enrolling residents aged 40-79 years across 12 southern states. The large majority (86%) of participants were enrolled at community health centers (CHCs), institutions providing basic and preventative health care in underserved communities, so the cohort includes low-income segments of society typically not included in large numbers in other cohorts. Both African and non-African American participants were included, with more than twice as many African American participants enrolled to help address under-representation of black people in previous epidemiologic studies of cancer. Study participants completed a detailed baseline questionnaire, via in-person computer-assisted interviews at CHCs, and nearly 90% provided a biologic specimen. Follow-up of the cohort and identification of cancer cases was conducted with national mortality registers and with linkage to state cancer registries.

**Swedish Mammography Cohort (SMC) and Swedish Men Cohort (SMC_COSM)**

The Swedish Mammography Cohort (SMC) and the Cohort of Swedish Men (COSM) are two large population-based prospective cohorts from central Sweden. The SMC was initiated between 1987 and 1990 when all women born in 1914-1948 and residing in Uppsala and Vaestmanland counties were invited; response rate 74%

(n=66,651). The COSM started in late 1997, with the invitation of all men born in 1918-1952 and residing in Vaestmanland and Ãrebro county; response rate 49% (n=48,850). Questionnaire data on diet and other lifestyle factors was collected at the start of the studies and has been updated repeatedly during follow-up. Further, biological samples (saliva, blood) have been collected with signed informed consent and are available for DNA extraction. The cohorts are annually matched to the Swedish Cancer Register for ascertainment of incident cancer cases.

**UK Biobank (UKB)**

The UK Biobank (UKB) is a large-scale, prospective cohort study established to improve the prevention, diagnosis, and treatment of a wide range of serious and life-threatening illnesses, including cancer, cardiovascular disease, and dementia. Between 2006 and 2010, over 500,000 men and women, aged 40-69 years, were recruited from across the United Kingdom. At baseline, participants attended one of 22 assessment centers where they provided detailed information via touchscreen questionnaires on their lifestyle, environment, and medical history. Physical measurements were also taken, and biological samples, including blood, urine, and saliva, were collected for long-term storage and analysis, from which extensive genetic and other biomarker data have been generated. Participant health outcomes are followed long-term through comprehensive electronic linkage to national health records, including hospital data and national cancer and death registries, allowing for robust ascertainment of incident diseases and mortality. CRC cases were defined as participants with primary invasive CRC diagnosed, or who died from CRC according to ICD9 (1530-1534, 1536-1541) or ICD10 (C180, C182-C189, C19, C20) codes.

**VITamins And Lifestyle (VITAL)**

The VITamins And Lifestyle (VITAL) cohort comprises of 77,721 Washington State men and women aged 50 to 76 years, recruited from 2000 to 2002 to investigate the association of supplement use and lifestyle factors with cancer risk. Participants were recruited by mail, from October 2000 to December 2002, using names purchased from a commercial mailing list. All participants competed a 24-page questionnaire and buccal-cell specimens for DNA were self-collected by 70% of the participants. Participants are followed for cancer by linkage to the western Washington SEER cancer registry and are censored when they move out of the area covered by the registry or at time of death.

**Women's Health Initiative (WHI)**

The Women's Health Initiative (WHI) is a long-term national health study focused on defining the risks and benefits of strategies to prevent heart disease, cancer, and osteoporotic fractures in postmenopausal women. From 1993 to 1998, the WHI recruited 161,808 post-menopausal women aged 50 to 79 at 40 clinical centers throughout the United States. WHI comprised a clinical trial arm, an observational study (OS) arm, and several extension studies. The clinical trials evaluated the effects of postmenopausal hormone therapy, a low-fat eating pattern, and calcium and vitamin D supplementation. At baseline, participants provided extensive information on medical history, lifestyle, and diet via questionnaires, and underwent physical measurements and blood sample collection. Health outcomes are ascertained through annual questionnaires and confirmed by review of medical records and linkage to the National Death Index.

#### Development and Description of Community Advisory Panel

##### Development of Community Advisory Panel

The Community Advisory Panel (CAP) was formed in 2021 to ensure the research considers the needs and preferences of the community members who may benefit from the results of the study. The CAP provides input on the approach and findings of the project from the community perspective, advises the research team on communication with local communities, provides feedback on risk tool development, content, and appearance, and recommends strategies to recruit participants to test the risk communication tool.

##### Description of CAP members and meeting agendas and discussions

To ensure a geographically and racially and ethnically diverse CAP, potential members were recruited from across U.S. with support from the Community Outreach and Engagement (COE) leaders of the four collaborating institutions: Fred Hutchinson Cancer Center (Washington), Moffitt Cancer Center (Florida), Cedars-Sinai Medical Center (California), and Cleveland Clinic (Ohio).

Individuals who expressed interest in participating were contacted via email and provided with a one-page CAP Charter which included a background, summary of the study, the intended role of the CAP, as well as time commitment and compensation. Those who agreed to participate were then asked to complete a brief online intake survey that provided the research team with contact information (e.g., phone number, email, preferred method of contact), occupation, demographics (age range, race, ethnicity), and open-ended questions about their prior involvement working with a CAP and what the individual was hoping to gain or learn more about as a result of this experience. There have been eight individuals serving on the CAP for this project.

The ACCELERATE CAP meetings occur semi-annually via teleconference and last 60-90 minutes. CAP members also receive an electronic research newsletter semi-annually with research updates and planning for the upcoming CAP meeting. CAP meetings provide bidirectional educational opportunity and engagement, where CAP members engage in facilitated discussion with the CAP research leads.

To prepare the CAP members for collaboration in the research project, the CAP research leads organized trainings on CRC, CRC screening, precision medicine, PRS, and cost-effectiveness to optimize screening. These interactive sessions established the foundation to equip the CAP to provide recommendations on the development of this risk tool. CAP meetings in 2024-2025 focused on gaining CAP member feedback and on development of the risk tool for this study. CAP members receive a $100 honorarium at the end of each calendar year for their participation in CAP meetings. On average, 71% of CAP members attended each meeting. About 50% of CAP meeting time was reserved for CAP member input, discussion, and questions.

#### Data Security and Approval

As advances in genomics continue to shape personalized medicine, it is imperative to ensure that the implementation of such tools adheres to rigorous standards to safeguard both the data and the individuals involved. Here we provide details about data security and ownership, compliance with regulatory standards, and user disclosures and acknowledgements required prior to tool use.

The Institutional Review Board (IRB) of Fred Hutchinson Cancer Center gave ethical approval for this work under file #3995.

**Data Security and Ownership:** The genetic risk prediction tool involves the use of individual genomic data provided by the users, along with a lifestyle questionnaire answered by the users in response to their lifestyle risk factors thereby necessitating the need to safeguard confidentiality and prevent unauthorized access. Encryption protocols, secure storage solutions, and access controls have been implemented to help mitigate potential security risks. Additionally, continuous monitoring and updates to security protocols are in place as part of Institutional Security policies. The genetic data transformation, quality control, imputation and polygenic risk score (PRS) calculation are performed in real-time, ensuring a dynamic and immediate analysis without the need to store or retain any individual-level data. The process occurs instantaneously without leaving a footprint, prioritizing privacy and data security by avoiding the storage of sensitive genetic and lifestyle information. This ephemeral approach (which can also be described as “on-the-fly” approach) reduces the likelihood of a security incident as calculations are generated on demand, serving the purpose of risk prediction without compromising the privacy of participants. Users retain ownership and control over their genetic and lifestyle data during ephemeral processing.

**User Acknowledgement:** Before uploading data and answering questions, all users must acknowledge review of the disclaimer that the site is not intended to provide medical advice and indicate acceptance of the terms and conditions regarding privacy.

**Compliance:** The risk prediction tool will adhere to industry standards for information security such as the NIST security framework. Security risks were assessed by the Fred Hutch Information Technology team. Legal and regulatory review was provided by the Fred Hutch Offices of Compliance, Regulatory Affairs and General Counsel.

**Questionnaires**

**PLEASE ANSWER THE FOLLOWING QUESTIONS TO GET STARTED**

BASIC INFORMATION & MEASUREMENTS

1. **What is your current weight**? (in pounds)

   **Current weight:** ___ ___ ___ pounds
   ________kg

2. **What is your current height?**

   **Current height:** ____ feet ____ inches

   ______cm

3. **What is your current age?**

   **Age:** ___ ___ years

4. **What sex were you assigned at birth?**
   - Female
   - Male
   - Rather not answer

   Note: By choosing 'Rather not answer', we will provide estimated risks of males and females, respectively.

NEXT: FAMILY HISTORY & DIET

5. **Have any of these members of your family had a colorectal cancer diagnosis? Biological parent, biological sibling, biological child. Biological means related by blood.**
   - Yes
   - No
   -
6. **Have you ever had a <u>colonoscopy in the last 10 years</u> or sigmoidoscopy? (A colonoscopy is a test that looks at the inside of your rectum and colon (large intestine)**

**using a long flexible tube with a light and small video camera on the end. Before the procedure, you are given a sedative (medication to help you relax). A sigmoidoscopy is a test that looks at the inside of your rectum and lower colon using a flexible lighted tube. This examination is usually done in a doctor's office. Most people don't need a sedative (medication to help them relax) for this test.)**

- Yes
- No
- Unknown

7. **On average, how often do you eat red meat? (This does NOT include chicken or fish.)** *A serving of red meat is: 2-3 ounces of red meat, about the size of a deck of cards. Red meats include beef, steak, hamburger, prime rib, veal, lamb, pork, bacon, and pork sausages.*
    - 10 or more servings per week
    - 8-9 servings per week
    - 7-8 servings per week
    - 4-6 servings per week
    - Less than 4 servings per week
    - None

8. **On average, how often do you eat a serving of fruit?** *A serving of fruit is: 1 medium fresh fruit, or ½ cup of chopped, cooked or canned fruit, or ¼ cup of dried fruit, or 6 ounces of fruit juice.*

    - 6 or more servings per day
    - 4-5 servings per day
    - 2-3 servings per day
    - One serving per day
    - Less than 1 serving per day

NEXT: HEALTH & LIFESTYLE

9. **In an average week, about how many alcoholic beverages do you drink?** *1 alcoholic beverage = 4-oz. glass of wine, or 12-oz. can or bottle of beer or hard cider, or 1-oz. serving of sake or liquor.*

    - 28 or more per week
    - 21 to 27 per week
    - 14 to 20 per week
    - 7 to 13 per week

- 1 to 6 per week
- Less than 1 per week
- None

10. **Have you smoked at least 100 cigarettes in your entire life?**

    - Yes *(Go to Question 11 and 12)*
    - No *(Go to Question 13)*

11. **During periods when you smoked regularly (or currently smoke), how many cigarettes did you typically smoke in a day?** *(1 pack = 20 cigarettes.)*

    |___|___|___| Number of cigarettes (per day)

12. **How many years did you smoke regularly?** If you currently smoke please include all years up to now.

    ___ ___ years

13. **Has a doctor ever told you that you had diabetes, also known as diabetes mellitus?** *(Note: this does not include diabetes you had only during pregnancy)*

    - Yes
    - No

NEXT: MEDICATIONS & DEMOGRAPHICS

**14. The following question is for women only : Have you ever used hormone replacement therapy (estrogen alone or estrogen plus progestin)?**

- Yes
- No

**15. Do you regularly take aspirin (at least twice a week for a month or longer)?**

- Yes

- No

**16. Do you regularly take NSAIDs such as ibuprofen, naproxen, or other non-steroidal anti-inflammatory drugs (at least twice a week for a month or longer)?**

- Yes
- No

**17. Do you regularly take calcium supplements (at least twice a week for a month or longer)?**

- Yes
- No

18. **Are you Latino/Hispanic?**

- Yes
- No (If you answer “No” go to Question 19)

19. **What is your race?** (*Check **one** that apply*)

- Asian
- Black or African American
- White
- Other or Mixed Race

Note: Unfortunately, we currently do not have sufficient data to develop risk prediction models for racial and ethnic groups not listed here. In those instances, we will use the average risk across the populations.

**Sample Reports**

- **PRS only (9 pages in total)**

**Fred Hutchinson Cancer Center**
MyGeneRisk Colon Assessment
Generated: January 28, 2026

Fred Hutch Cancer Center

### Your MyGeneRisk Colon Report

**When to Start Screening**

Schedule your screening now. It is important to start screening now if you haven't already.

**What to Do Now**

Schedule your colorectal cancer screening with your doctor. Regular screening is the best way to prevent colorectal cancer because it can find and remove polyps (small bumps of tissue in your colon) before they turn into cancer. Don't wait—screening is an important step to protect your health.

#### Your Colorectal Cancer Risk for the Next 10 Years

| Average Risk | Your Risk: Above Average Risk | High Risk |
|---|---|---|
| Your chance of getting colorectal cancer is about the same as most people in the US. | Your chance of getting colorectal cancer is a bit higher than most people in the US. | Your chance of getting colorectal cancer is as high as someone whose close blood relative, like a parent or sibling, had the disease. |

**Your estimated risk of developing colorectal cancer in the next 10 years = 1.51%**

#### Your Colorectal Cancer Risk for the Next 10 Years

**What this means:** If we look at **100** people with the same age and sex as you, we expect about **1.5** of them to develop colorectal cancer in the next 10 years.

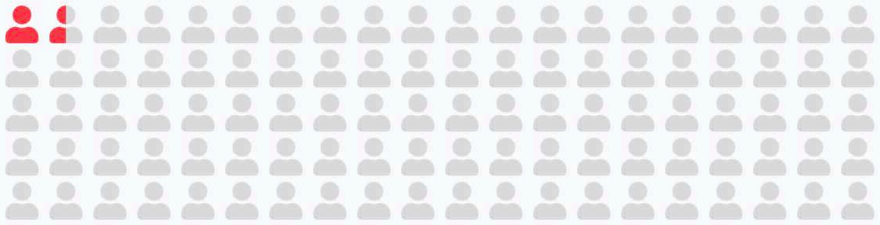

#### Your Lifetime Colorectal Cancer Risk

This graph shows how your chances of getting cancer during your lifetime compared to the average person of your age and sex assigned at birth.

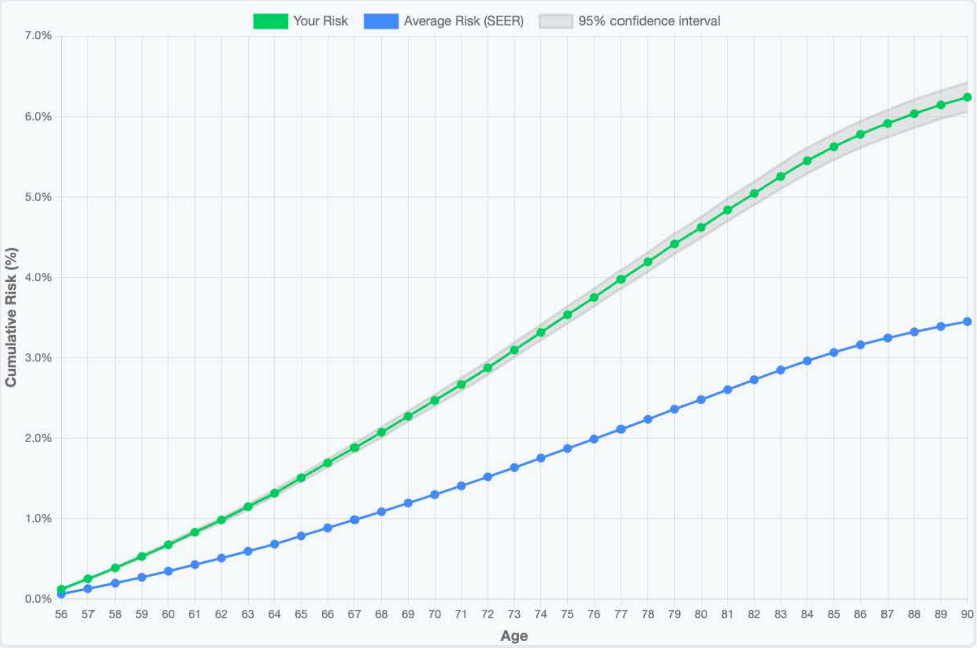


- The blue line shows the average lifetime risk for someone your age based on national data (details see: https://seer.cancer.gov/about/).
- The green line shows your personal risk, which may be higher or lower depending on various factors like your genes and lifestyle.
- If the green line is above the blue line, your personal risk for colorectal cancer is HIGHER than average. This means you are more at risk for developing colorectal cancer than the average person.
- The gray shaded area around the green line is called a confidence interval — it means while we provide the best estimate of your risk, we're not 100% sure of your exact risk, but that gray area has a 95% chance that covers your exact risk.

#### What Should I Do Next?

Understanding your risk is the first step. Here's what we recommend based on your results.

**Schedule your screening today!**

Everyone 45 and older, even if there are no risk factors, should get regular colorectal cancer screenings. If someone in your close family (like a parent or sibling) has colorectal cancer, you should start before age 45.

Screening is free as a preventive service covered by the Affordable Care Act.

---

**How to get screened:**

Contact your doctor or other health care provider. Tell them you'd like to get screened for colorectal cancer by either getting a

- Fecal immunological test (FIT) kit or
- Colonoscopy.

Take this report with you and review it with your doctor or other healthcare provider.

#### What Should I Do Next?

**Screening Tests for Colorectal Cancer**

Two common screening tests for colorectal cancer are a FIT test and a colonoscopy. Talk with your doctor or other health care provider to decide which test is best for you.

##### FIT (Fecal Immunological Test)

1 time every year (age 45-75)

**What it is:**

An easy test for colorectal cancer that you do at home.

**How it works:**

1. **Get a kit:**
   Ask your doctor to give or send you one.
2. **Collect a sample:**
   Use the stick in the kit to get a small sample of your poop.
3. **Mail it back:**
   Send the sample to the lab using the envelope in the kit.

**The results and next steps:**

Your results will either be "normal" or "abnormal"

**Normal test?** Retake it in 1-3 years

**Abnormal test?** You will need to schedule a colonoscopy so a doctor can look at your colon and learn more about what's going on.

**The benefits:**

- You can do this test at home, at your convenience.
- A FIT can be combined with stool DNA testing, like Cologuard (you send in one poop sample for both tests).

**The limits of FIT screening:**

FIT looks for blood in your poop. It does not look at the inside of your colon and rectum.

#### What Should I Do Next?

**Screening Tests for Colorectal Cancer**

##### Colonoscopy

Every 10 years (age 45-75)*

**How it works:**

1. **Schedule your colonoscopy:**
   Your regular doctor or other health care provider will refer you to a special doctor who does colonoscopies.
2. **Do bowel prep:**
   Before the colonoscopy, you will follow a special diet to clean your colon of any poop.
3. **Go to the hospital:**
   Most colonoscopies are done at a hospital.
4. **Get a sedative:**
   You'll be given medicine to help you relax and be comfortable (sedative).
5. **Doctor does the colonoscopy:**
   The physician will put a thin, flexible tube called a colonoscope into your rectum to look inside. They look for polyps (which may or may not be cancer), tumors, and other abnormal tissue. If the doctor finds anything, they can remove it and send it to a lab for testing (biopsy) to see if the cells are cancer.

**The results and next steps:**

Usually, right after your colonoscopy, the doctor meets with you. They share what they saw, and if they removed polyps or took a biopsy. If a biopsy is done, they will tell you when to expect results.

Most people get a colonoscopy every 10 years from age 45 to 75. If you have an increased risk of colorectal cancer, talk with your doctor about how often you should get a colonoscopy.

**The benefits:**

The doctor:

- Gets an in-depth look at your colon and rectum
- Can remove any polyps (growths that can become cancer over time)
- Can take a biopsy if they need to. A biopsy is when a small piece of tissue is removed and examined in a lab.

**Earlier and more frequent screening is necessary for those at increased risk. Please consult your doctor.*

#### Reduce Your Risk Through Healthy Habits

These evidence-based lifestyle changes can help lower your colorectal cancer risk.

##### Eat a Healthy Diet

You can lower your risk of cancer by eating a diet high in fiber and plant-based foods. Fiber helps your body digest food and keep your gut bacteria healthy. Plant-based foods give you important nutrients that help lower inflammation in the body and protect against cancer.

**Eat plenty of:**

- Vegetables: 2.5–3 cups per day
- Fruit: 1.5–2 cups per day
- Whole grains: at least 3 servings per day
  - Examples: brown rice, whole-wheat breads & pasta, oats, quinoa, and more
  - Benefit: increases your fiber intake, an important protective factor against cancer

**Limit intake of:**

- Red meat (beef, pork, lamb)
- Processed meats (hot dogs, sausage, deli meats)

##### Be Active

Regular exercise keeps your heart strong, your brain sharp, and your mood more positive, while lowering your risk of serious diseases like heart disease, diabetes, and some cancers including colorectal cancer.

**Recommendation:**

- Move your body for at least 30 minutes each day. Find things you enjoy doing, like taking a walk, dancing, jogging, playing sports, practicing yoga, weightlifting, or other activities.

##### Maintain a Healthy Weight

Being overweight increases your risk of colorectal cancer because extra body fat can trigger inflammation, which may help cancer grow and spread.

**Recommendation:**

- Eat a healthy diet and move your body every day to help maintain a healthy weight.

##### Quit Smoking

People who smoke are more likely to develop a higher number of colon polyps, and their polyps tend to be larger. These polyps can turn into cancer over time.

**Recommendation:**

- Try not to smoke and avoid secondhand smoke.

##### Limit Alcohol Intake

Drinking alcohol can keep your body from getting the nutrients it needs to stay healthy. This can make it easier for cancer to grow.

**Recommendation:**

- To help prevent cancer, it's best not to drink alcohol. If you do drink, limit alcohol to no more than 2 drinks per day for men, and 1 drink per day for women.

#### Your Answers

Review the information you provided during your assessment. If any answers seem incorrect, you may want to retake the assessment or discuss with your healthcare provider.

**Sex assigned at birth**
Male

**Age**
55 years old

**Are you Latino/Hispanic?**
No

**Race**
White

#### About this risk prediction tool

To estimate an individual's risk of developing colorectal cancer, we developed a prediction tool that uses both genetic information and lifestyle factors. The genetic part, called polygenic risk score (PRS), adds up the effects of over 1 million genetic variants associated with colorectal cancer risk. These scores were developed using data from large studies of Asian, Black or African American, Hispanic, and non-Hispanic White populations and their effect sizes were estimated accordingly. The lifestyle factors include height, body weight, history of type 2 diabetes, smoking, alcohol use, use of regular aspirin or nonsteroidal anti-inflammatory drugs, postmenopausal hormone use, intake of calcium supplements, red meat, fruit, history of colonoscopy or sigmoidoscopy, and family history of colorectal cancer. We brought together data on these factors from multiple racially and ethnically diverse cohorts and estimated their effect sizes using pooled individual-level data. Using these effect size estimates for PRS and lifestyle factors, we calculated an individual's chance of developing colorectal cancer over specified time periods (here, 10 years or lifetime) given their PRS and lifestyle choices, while also considering the risk of death from other causes.

This tool does not test for rare, high-risk genes that carry mutations for Lynch syndrome, familial adenomatous polyposis or others. If you have a family history of colorectal cancer, talk with your doctor about a screening plan that includes genetic testing.

##### Disclaimer

MyGeneRisk Colon was developed by a team led by Drs. Ulrike (Riki) Peters and Li Hsu at Fred Hutch Cancer Center, with support from National Institutes of Health (R01-CA244588, P50-CA285275, and R01CA297681). The risk estimates in this tool are based on genetic and lifestyle data from a large number of colorectal cancer studies.

This tool is meant for informational and educational purposes only. It does not diagnose, treat, or prevent any disease, including cancer. The risk estimates provided are based only on the information you share.

The results should not be used to replace professional medical care and advice. If you have received a positive screening test for colorectal cancer or have concerns about your health, contact a qualified healthcare provider as soon as possible.

Fred Hutch cannot control how data is used by others. Please refer to the privacy policies and terms of use when you share your personal data with any third parties.

**Fred Hutchinson Cancer Center**

1100 Fairview Ave N
Seattle, WA 98109

**Questions?**

Visit MyGeneRisk-Colon.FredHutch.org

*© 2026 Fred Hutchinson Cancer Center. All rights reserved.*

- **Lifestyle only (page 1-3 and 8, the remaining pages are same as the PRS risk report)**

**Fred Hutchinson Cancer Center**
MyGeneRisk Colon Assessment

Generated: January 28, 2026

Fred Hutch Cancer Center

### Your MyGeneRisk Colon Report

##### When to Start Screening

Schedule Your screening now. It is important to start screening now if you haven't already.

##### What to Do Now

Schedule your colorectal cancer screening with your doctor. Regular screening is the best way to prevent colorectal cancer because it can find and remove polyps (small bumps of tissue in your colon) before they turn into cancer. Don't wait—screening is an important step to protect your health.

#### Your Colorectal Cancer Risk for the Next 10 Years

Your Risk

**Average Risk**

Your chance of getting colorectal cancer is about the same as most people in the US.

**Above Average Risk**

Your chance of getting colorectal cancer is a bit higher than most people in the US.

**High Risk**

Your chance of getting colorectal cancer is as high as someone whose close blood relative, like a parent or sibling, had the disease.

**Your estimated risk of developing colorectal cancer in the next 10 years = 0.50%**

#### Your Colorectal Cancer Risk for the Next 10 Years

**What this means:** If we look at **100** people with the same age and sex as you, we expect about **0.5** of them to develop colorectal cancer in the next 10 years.

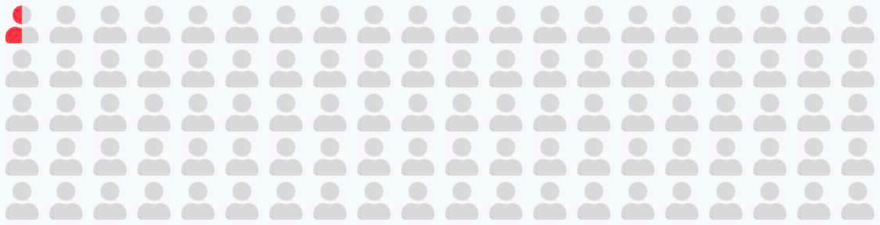

#### Your Lifetime Colorectal Cancer Risk

This graph shows how your chances of getting cancer during your lifetime compared to the average person of your age and sex assigned at birth.

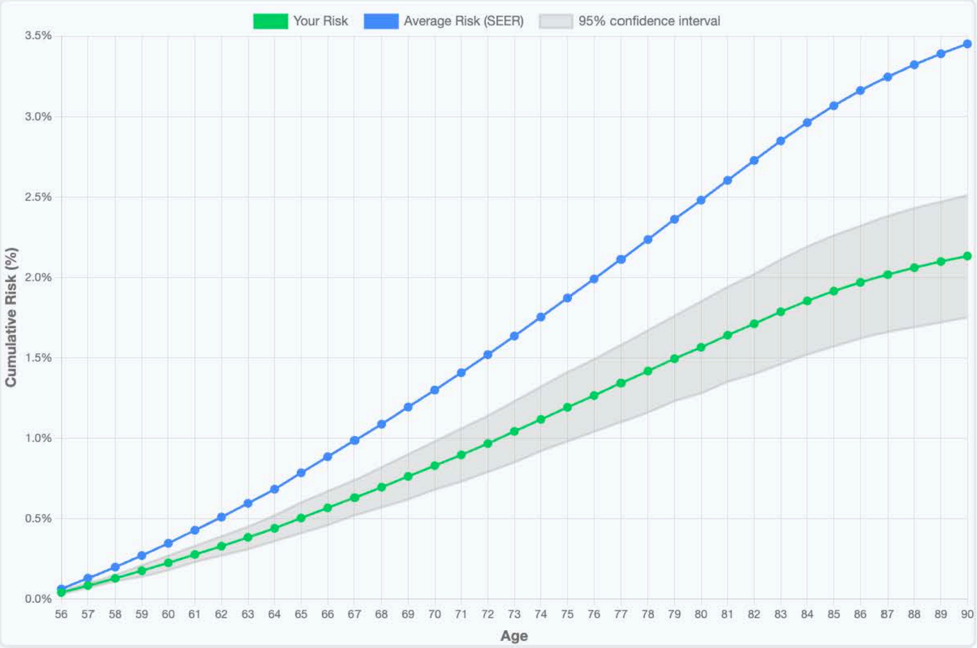


- The blue line shows the average lifetime risk for someone your age based on national data (details see: https://seer.cancer.gov/about/).
- The green line shows your personal risk, which may be higher or lower depending on various factors like your genes and lifestyle.
- If the green line is above the blue line, your personal risk for colorectal cancer is HIGHER than average. This means you are more at risk for developing colorectal cancer than the average person.
- The gray shaded area around the green line is called a confidence interval — it means while we provide the best estimate of your risk, we're not 100% sure of your exact risk, but that gray area has a 95% chance that covers your exact risk.

##### Limit Alcohol Intake

Drinking alcohol can keep your body from getting the nutrients it needs to stay healthy. This can make it easier for cancer to grow.

**Recommendation:**

- To help prevent cancer, it's best not to drink alcohol. If you do drink, limit alcohol to no more than 2 drinks per day for men, and 1 drink per day for women.

#### Your Answers

Review the information you provided during your assessment. If any answers seem incorrect, you may want to retake the assessment or discuss with your healthcare provider.

**Sex assigned at birth**
Male

**Age**
55 years old

**Are you Latino/Hispanic?**
No

**Race**
White

**Weight**
181 lbs

**Height**
5'11"

**Family history of colorectal cancer**
No

**Previous colonoscopy or sigmoidoscopy**
Yes

**Red meat consumption**
4-6 servings per week

**Fruit consumption**
One serving per day

**Alcoholic beverages per week**
1 to 6 per week

**History of diabetes**
No

**Smoked at least 100 ciga-rettes lifetime**
No

**Regular aspirin use**
No

**NSAID use (ibuprofen, Advil, etc.)**
No

**Calcium supplement use**
Yes

- **PRS and lifestyle combined (page 1-3 and 8, the remaining pages are same as the PRS risk report)**

**Fred Hutchinson Cancer Center**
MyGeneRisk Colon Assessment
Generated: January 28, 2026

Fred Hutch
Cancer Center

#### Your MyGeneRisk Colon Report

**When to Start Screening**

Schedule your screening now. It is important to start screening now if you haven't already.

**What to Do Now**

Schedule your colorectal cancer screening with your doctor. Regular screening is the best way to prevent colorectal cancer because it can find and remove polyps (small bumps of tissue in your colon) before they turn into cancer. Don't wait—screening is an important step to protect your health.

##### Your Colorectal Cancer Risk for the Next 10 Years

| Average Risk | Your Risk: Above Average Risk | High Risk |
| --- | --- | --- |
| Your chance of getting colorec-tal cancer is about the same as most people in the US. | Your chance of getting col-orectal cancer is a bit higher than most people in the US. | Your chance of getting colorec-tal cancer is as high as some-one whose close blood relative, like a parent or sibling, had the disease. |

**Your estimated risk of developing colorectal cancer in the next 10 years = 0.87%**

#### Your Colorectal Cancer Risk for the Next 10 Years

**What this means:** If we look at **100** people with the same age and sex as you, we expect about **0.9** of them to develop colorectal cancer in the next 10 years.

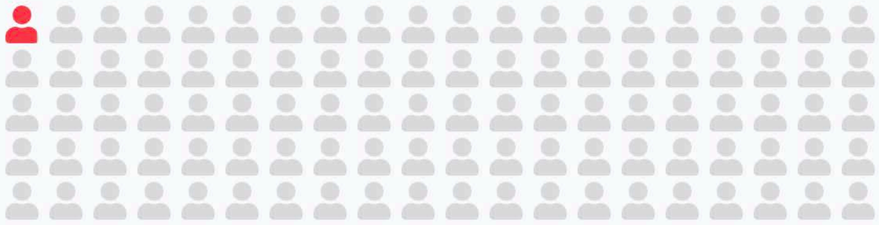

#### Your Lifetime Colorectal Cancer Risk

This graph shows how your chances of getting cancer during your lifetime compared to the average person of your age and sex assigned at birth.

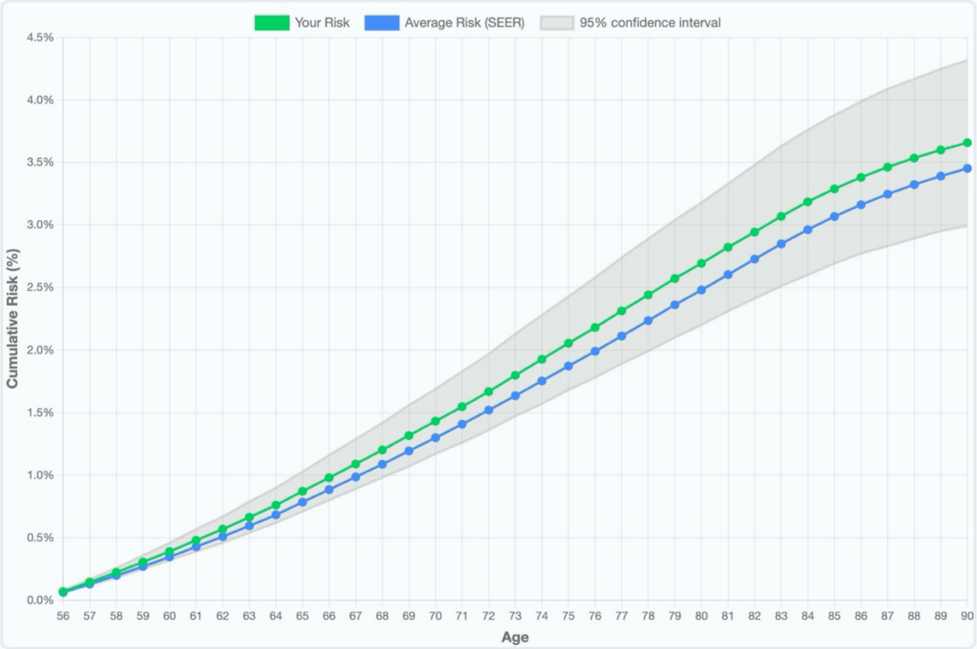


• The blue line shows the average lifetime risk for someone your age based on national data (details see: https://seer.cancer.gov/about/).

• The green line shows your personal risk, which may be higher or lower depending on various factors like your genes and lifestyle.

• If the green line is above the blue line, your personal risk for colorectal cancer is HIGHER than average. This means you are more at risk for developing colorectal cancer than the average person.

• The gray shaded area around the green line is called a confidence interval — it means while we provide the best estimate of your risk, we're not 100% sure of your exact risk, but that gray area has a 95% chance that covers your exact risk.

##### Limit Alcohol Intake

Drinking alcohol can keep your body from getting the nutrients it needs to stay healthy. This can make it easier for cancer to grow.

**Recommendation:**

- To help prevent cancer, it's best not to drink alcohol. If you do drink, limit alcohol to no more than 2 drinks per day for men, and 1 drink per day for women.

#### Your Answers

Review the information you provided during your assessment. If any answers seem incorrect, you may want to retake the assessment or discuss with your healthcare provider.

**Sex assigned at birth**
Male

**Age**
55 years old

**Are you Latino/Hispanic?**
No

**Race**
White

**Weight**
185 lbs

**Height**
5'11"

**Family history of colorectal cancer**
No

**Previous colonoscopy or sigmoidoscopy**
Yes

**Red meat consumption**
4-6 servings per week

**Fruit consumption**
One serving per day

**Alcoholic beverages per week**
1 to 6 per week

**History of diabetes**
No

**Smoked at least 100 cigarettes lifetime**
No

**Regular aspirin use**
No

**NSAID use (ibuprofen, Advil, etc.)**
No

**Calcium supplement use**
Yes

**Polygenic Risk Score (PRS) Pipeline and QC Checks**

Genotype data from direct-to-consumer (DTC) genotyping providers are accepted in all major plain-text formats (e.g., CSV, TSV, TXT) containing variant-level information. Each file undergoes quality control (QC) performed within the user's browser to ensure structural integrity, completeness, and compatibility with downstream PRS computation.

The following QC steps involve:

1. Input structure and genome coverage: Verification that all autosomes (chromosomes 1–22) are present. Each chromosome is required to contain a minimum of 100 SNPs per megabase (Mb) to ensure adequate marker density.
2. Variant count threshold: The total number of SNPs exceeds 500,000.
3. Missing call rate: The missing genotype call rate is <=5%.
4. Column integrity: Presence of the four required columns: rsid, chromosome, position, and genotype.
5. Genome build: If a header is present, it is required to reference one of the supported genome assemblies GRCh36, GRCh37, or GRCh38. Files lacking this information are flagged for genome build inference during preprocessing.

Only files that meet all QC criteria are split by chromosome, uploaded to independent AWS Lambda functions, and advanced to the preprocessing stage. Each Lambda instance:

1. Converts chromosome-level genotyping data to Variant Call Format (VCF).
2. Removes sites with missing genotyping calls, structural variants and insertions/deletions.
3. Determines genome build: if genome build is listed in the header, the provided assembly information is used. Otherwise, genome build is inferred by matching the provided dbSNP ids and SNP locus information (chromosome and position) to dbSNP databases from multiple assemblies, and the best match is selected, or fails if no database shows at least a 50% match.
4. Lifts over variants to reference build GRCh37 when necessary and aligns them to the plus strand.
5. Phases variants using Eagle2 to infer haplotype structure
6. Imputes missing genotypes using Minimac4 with the 1000 Genomes Phase 3 reference panel.

The output contains dosage value for each variant representing the expected allele count on a continuous scale from 0-2, where 0 corresponds to the homozygous reference genotype, 1 to the heterozygous genotype, and 2 to the homozygous alternative genotype. The dosages are used to:

1. calculate a raw Polygenic Risk Score (PRS) for each chromosome as the sum of imputed genotype dosages weighted by the variant effect sizes:

   $$PRS = \sum_i \beta_i \times dosage_i$$

   where $\beta_i$ denotes the effect size for variant $i$.

2. Compute principal components (PCs) using pre-calculated loadings derived from the 1000 Genomes reference dataset.

Each Lambda instance returns the raw PRSs and PCs back to the client, after which all temporary files are deleted from the AWS Lambda instances.

The client browser collects the PRSs and PCs from all 22 chromosomes, combines chromosome-specific PRS values into a genome-wide PRS, and recalibrates the overall score using the PCs to adjust for ancestry-related variation. The recalibrated PRS is then combined with participant-reported lifestyle and demographic information to estimate 10-year and lifetime colorectal cancer risk.

The estimated risk is compared against age- and sex-specific incidence rates obtained from the Surveillance, Epidemiology, and End Results (SEER) program.

The browser then generates a personalized risk report with intuitive graphical displays and textual explanations. The report also includes recommendations for modifiable risk factors, such as diet, alcohol consumption, and physical activity, which may reduce colorectal cancer risk. Finally, a summary of the tool for healthcare providers is also provided.

**Evaluation of Single-Sample Imputation Pipeline**

Imputation protocols typically perform joint phasing and imputation in batches, leveraging shared haplotype structure. However, this approach is impractical in the current setting, where individual-level data arrive sequentially, and PRSs must be calculated under time constraint. Single-sample imputation is the approach to go.

To ensure dosage accuracy from single-sample imputation, we performed validation using 830 samples from the Personal Genome Project (PGP).[1] Figure S3 shows the first 2 PCs of these samples along with 2,504 reference samples from the 1000 Genomes Project (661 African, 347 Admixed American, 504 East Asian, 503 European, and 489 South Asian). As self-reported race and ethnicity were not available for these PGP samples, we assigned ancestral groups based on population structure in principal component space. Specifically, we applied k-means clustering (k = 5) to the study samples using the first two principal components. Each cluster was then assigned to one of the ancestry groups defined by the 1000 Genomes Project by computing the Euclidean distance from its centroid to each ancestry-group centroid and selecting the group with the minimum distance. This results in the following assignments: 6 African, 265 Admixed American, 36 East Asian, 523 European, 25 South Asian.

Specifically, each of the 830 samples was imputed twice:

- Single-sample workflow: Each genome was individually imputed using the MyGeneRisk pipeline, aligning and processing each sample independently against the 1000 Genomes Phase 3 reference panel.
- Batch workflow: The same preprocessed genotype files were merged into a single VCF, then phased jointly using Eagle2 and imputed with Minimac4 using the same 1000 Genomes reference panel.

For each sample, PRS values were computed separately from the single-sample and batch-imputed dosages using the same SNP weights and reference calibration. The raw PRS values from the two imputation methods showed strong concordance (Figure S4; Pearson correlation coefficient = 0.98), which remained high after ancestry recalibration (Pearson correlation coefficient = 0.98).

While the overall correlation was strong, a subset of samples exhibited greater discordance between the two imputation methods, defined as $\left|PRS_{Single} - PRS_{batch}\right| > 0.2$. Samples of Admixed American ancestry showed markedly higher discordance rates of 60/265 (~22%), compared to 7/523 (~ 1.3%)

among European, 0/6 (0%) in African, 0/25 (0%) in South Asian and 1/11 (~9.1%) among East Asian ancestry samples. An investigation revealed a significant association between discordance and elevated heterozygosity rate across autosomes, which are consistent with individuals of recently admixed ancestries. These findings suggest that single-sample imputation may perform less accurately for individuals from admixed populations, possibly due to incomplete haplotype representation in the 1000 Genomes reference panel. In contrast, batch imputation benefits from joint phasing, which captures local haplotype sharing across diverse samples and better models mixed ancestry segments.

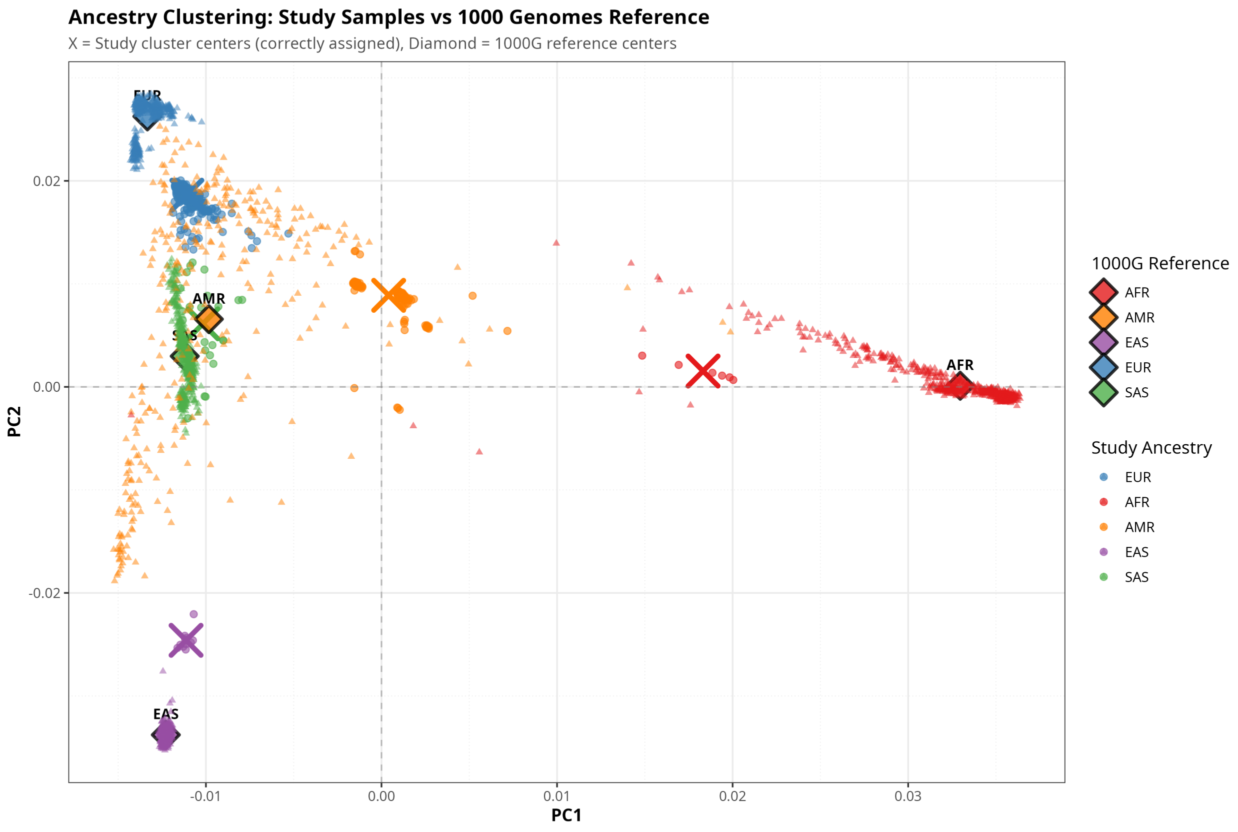


**Figure S3:** Scatterplot of first two principal components (PCs) for 830 samples from personal genome project (circles), along with 1000 Genomes Project Reference Panel (triangles). Diamonds indicate 1000 Genomes population centroids. AFR: African, AMR: Admixed American, EAS: East Asian, EUR: European, SAS: South Asian.

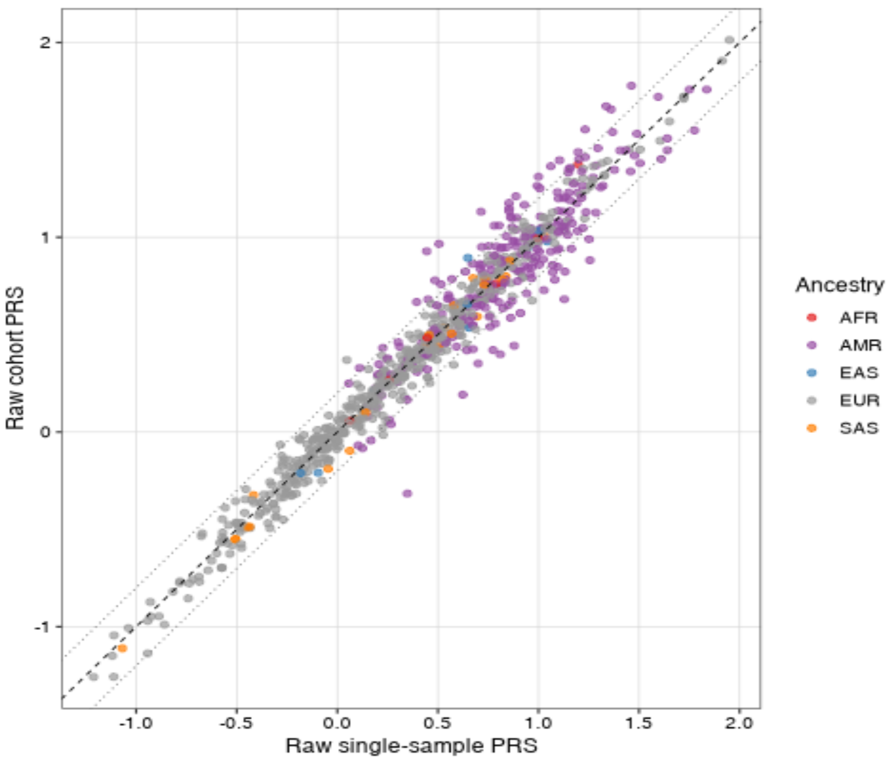


**Figure S4. Scatterplot of raw polygenic risk scores (PRS) from single-sample imputation (X-axis) and batch/cohort imputation (Y-axis). The** PRS scores were highly correlated (Pearson correlation coefficient =0.98) across inferred ancestry groups, including East Asian (EAS), South Asian (SAS), African (AFR), Admixed American (AMR), and European (EUR). Dotted lines indicate threshold for discordant samples (absolute distance > 0.2).
